# A feature ranking algorithm for clustering medical data

**DOI:** 10.1101/2023.09.30.23296349

**Authors:** Eran Shpigelman, Ron Shamir

## Abstract

**Objective:** Clustering methods are often applied to electronic medical records (EMR) for various objectives, including the discovery of previously unrecognized disease subtypes. The abundance and redundancy of information in EMR data raises the need to rank the features by their relevance to clustering.

**Methods:** Here we propose FRIGATE, an ensemble feature ranking algorithm for clustering. FRIGATE ranks the features by solving multiple clustering problems on subgroups of features, using game-theoretic principles to rank and weigh features. In every such problem, a Shapley-like framework is utilized to rank a selected set of features. In another version of the algorithm, multiplicative weights are employed to reduce the randomness in feature set selection. The code for the algorithms is available in: https://github.com/Shamir-Lab/FRIGATE.

**Results:** On simulated data and on eleven real genomics and EMR datasets, FRIGATE outperforms extant ensemble ranking algorithms, in solution quality and in speed.

**Conclusion:** Frigate can improve disease understanding by enabling better subtype discovery from EMR data.

## 1. Introduction

Due to the recent digitization revolution in medical data [1],vast amounts of patient medical information are now stored electronically, enabling a transformation of medical research. The number of clinical datasets available to researchers is growing [2].Some of them span a large range of clinical data types [3],and some even offer both genomic and medical information [4]. At the same time, these data have challenging characteristics, including data volume and numerous features. For example, the MIMIC-III dataset covers 46,520 patients, with 753 different lab tests and 14,567 different ICD-9 codes [3](each test or code is called a *feature*).

A variety of machine-learning studies have attempted to analyze medical data [5, 6]. Clustering, a fundamental unsupervised machine-learning approach, has been used for the discovery of new subtypes of known diseases [7, 8, 9, 10]. In the clustering process, a group of patients with feature value vectors are partitioned into subgroups based on the similarity of their features.

A key challenge in medical research is the interpretability of the results. Having found clusters in the data, we want to understand the most important features that distinguish them, in order to assign a clinical meaning to each cluster and obtain clinical insights. Dimension reduction methods [11]can handle datasets with many features, but they obscure the individual features. Here we aim to develop an algorithm that ranks the features according to their importance to the clustering task.

In recent years several ensemble feature ranking (EFR) algorithms were suggested. They create multiple subproblems induced by different subsets of features, cluster each of them and then use the ensemble of clustering solutions to evaluate the contribution of each feature. The EFR methods FRMV [12],FRCM [13],and FRSD [14]were shown to outperform the traditional filter and wrapper methods, including on medical data. In addition to ranking the full set of features, such methods can also be used for choosing a subset of important features [15].

We introduce here a new EFR algorithm called FRIGATE (Feature Ranking In clustering using GAme ThEory), which uses two concepts from game theory. The first is Shapley value, a measure of the contribution of every player to the group in a cooperative game [16, 17]. In our case the players are the features and the “game” is clustering. Shapley values are widely used for feature evaluation in classification [18, 17]but so far were not used in unsupervised learning for feature selection or ranking. The second is Multiplicative Weights (MW) [19],a framework to improve the selection of players by iteratively choosing the players from a distribution based on their performance so far. In FRIGATE we use MW to inform our selection of features for each clustering solution, thereby reducing the odds of choosing features that have been demonstrated to be unimportant. All extant ensemble algorithms choose feature subsets at random. To the best of our knowledge this is the first time that MW is used in feature selection for clustering.

The paper is organized as follows: In Section 2we provide brief relevant background on clustering, EFR for clustering and game theory concepts. In Section 3 we present the FRIGATE algorithm in two variants, describe the construction of simulated data, and introduce a novel approach to score rankings. In Section 4we measure the performance of FRIGATE and the extant EFR algorithms on simulated data and on eleven real genomics and EMR datasets. We conclude with a discussion of the results.

**Statement of Significance**

**Problem** Medical datasets encompass numerous variables, making it challenging to cluster patients and interpret the results effectively. A method to rank the features for clustering is needed.

**What is already known** Among the extant algorithms, ensemble methods have shown to be superior. However, no code was provided for them.

**What this Paper Adds** We present FRIGATE, a novel feature ranking for clustering algorithm for medical data. FRIGATE outperformed previously suggested algorithms. We also present two new evaluation metrics for comparing the methods. We provide the code of FRIGATE and the other tested algorithms.

## 2. Background

### Clustering

In the clustering problem the input is a set of points *x*_1_, …, *x*_*m*_ ∈ ℝ^*n*^ where *x*_*i*_ = (*x*_*i*1_, …, *x*_*in*_) and the goal is to partition them into groups called *clusters* so that points within each cluster are similar and points in different clusters are dissimilar. Similarity is measured by the squared Euclidean distance, i.e., 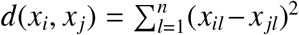. In our context, the points are the patients and the coordinates are the feature values. A fundamental, broadly used clustering heuristic is k-means [20].Given the number of clusters *k*, it selects points *y*_1_, … *y*_*k*_ ∈ ℝ*<*^*n*^ called centroids, assigns samples to the closest centroid and for each resulting set recomputes its center of mass as the new centroid. The solution score is 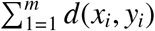. The process is iterated until convergence or for a preset number of iterations. k-modes [21]is a variant of k-means for *categorical data*, in which feature values are discrete (two or more) categories. The Hamming distance is used as the distance metric instead of Euclidean distance. k-prototypes [22]is an algorithm that clusters *mixed data*, i.e. data with both continuous and categorical features. The distance metric is:

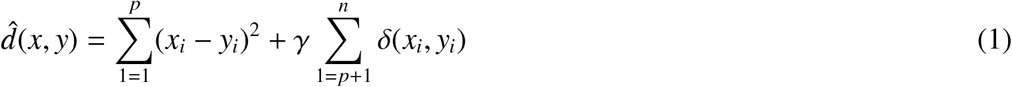

Where *x*_1_, … *x*_*p*_ are numerical variables, *x*_*p*+1_, … *x*_*n*_ are categorical variables, and Δ is the Hamming distance function. The γ factor determines the relative contribution of the categorical features. k-prototypes was one of the best performers in a recent mixed-data clustering benchmark [23].

### Ensemble feature ranking algorithms for clustering

In *feature ranking for clustering* the goal is to rank the features of a clustering problem by their contribution to the clustering solution. In ensemble feature ranking (EFR) algorithms, the problem is addressed by solving a set of clustering problems, each defined by restricting the input data to a subset of the coordinates, ranking the features in each solution, and aggregating the results into a final ranking. Several EFR algorithms were proposed, and they differ in the way they rank features per solution and in the aggregation method. In the FRMV algorithm [12],features in each solution are ranked based on some relevance measure (e.g. linear correlation), and the final feature ranking is done according to the average rank. FRCM [13],originally designed for genomic data, features in each solution are ranked based on a measure similar to Adjusted Rand Index [24].In FRSD [14],features ranking is based on the change in the silhouette score [25]after shuffling the values of the feature. In the FRCM and FRSD, instead of providing *k* as input, a prescribed range of *k* values is tested and the final score is based on the averaging the ranks.

### Shapley Values

In cooperative game theory, a set *N* of players can form coalitions. Each coalition *S* ⊆ *N* has a value *g*(*S*). According to the Shapley theory [16]the contribution of player *i* to group *S* ∪ {*i*} is defined as:

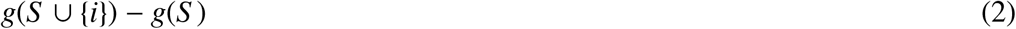

and the Shapley value of player *i* is a weighted average of its contributions over all possible *S* -s, i.e.:

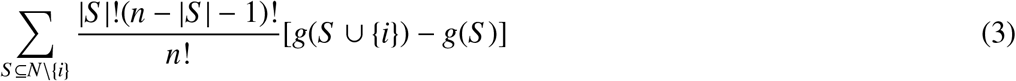

This value is widely used in supervised learning to measure the contribution of a feature to a prediction model [17].

### Multiplicative Weights

MW is an algorithmic update method used in game theory and algorithm design. The motivation of MW [19]is to iteratively improve the decisions one makes by gradually favoring decisions that were proven to be good so far. In our case the decisions are the features selected. The update is done by the Hedge rule [19]:

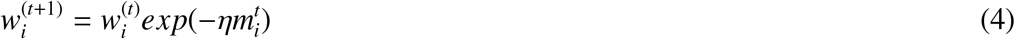

Where 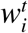 is the weight of feature *i* at the *t*-th iteration, η ≤ 1 is a parameter and 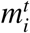 is the cost of feature *i* at iteration *t*. 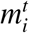 is a value in the range [−1, 1] that reflects how good decision *i* was in iteration *t*, where higher positive values correspond to worse decisions and negative values correspond to good decisions. A common practice, also used in our implementation, is to use non-negative values only.

## 3. Methods

### 3.1 The Frigate algorithm

FRIGATE is a new EFR algorithm inspired by the Shapley value. It performs multiple runs of k-means (or k-prototypes) on problems defined by feature subsets, and uses their clustering solution scores for computing feature importance. Here coalitions correspond to feature subsets, and the value of each subset *S* is its solution score.

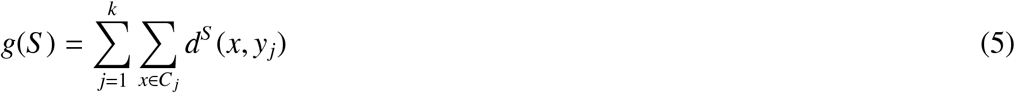

Here *k* is the number of clusters, *C* = {*C*_1_, …, *C*_*k*_} is the clustering solution, where *C*_*j*_ is the set of samples included in cluster *j* and *y*_*j*_ is the centroid of *C*_*j*_. *d*^*S*^ is the distance function on the sample vectors restricted to the coordinates in *S* . *d* is the squared Euclidean if all features are continuous, or the distance function (1)if features are mixed. Note that *g*(*S*) depends on both *S* and *C*. We call *g*(*S*) the *solution score*.

In the computation of feature importance, we deviate from equation (3). We define the *contribution* of feature *i* to the problem restricted to feature subset *S* as the difference between its solution score and the score obtained by the same clustering when the values of feature *i* are randomly shuffled among the samples, keeping the values of the rest of the features unchanged. This mimics the effect of removing that feature, while keeping the total number of features unchanged, in order to avoid normalization problems. The underlying idea is that disrupting a more important feature would lead to a larger decrease in the score.

Algorithm 1 (Algorithm 1)presents the procedure for continuous features. In iteration *t*, the algorithm selects at random a subset of features *h* and performs k-means on the corresponding submatrix *A*^(*t*)^. Once a solution has been obtained, we calculate the contribution of each feature *i* in *h*. For the final ranking we use the average scores of the features. See Supplementary 2 for a graphical demonstration of FRIGATE.

Note that FRSD can also be seen as a Shapley-like algorithm with a function *g* that uses the silhouette. However, a main difference is that FRIGATE does not rank the features on every iteration and accumulates the ranks for the final score, as in FRSD and FRMV, but instead summarizes the raw scores. That way poor clustering solutions that are based on non-informative features will have large *g*(*h*) values (line 8 in Algorithm 1) as well as large *g*_*v*_ values (line 12). This will limit the ability of these features to receive high scores, as they are calculated by subtracting the distance after shuffling the values of a feature from the original distance (line 13). Thanks to these properties of *g*(*h*) and *g*_*v*_, we do not need to use an additional factor, as FRSD does with silhouette, to assess the quality of the clusters. It also reduces the number of calculations and improves efficiency.

#### Algorithm 1

The FRIGATE Algorithm

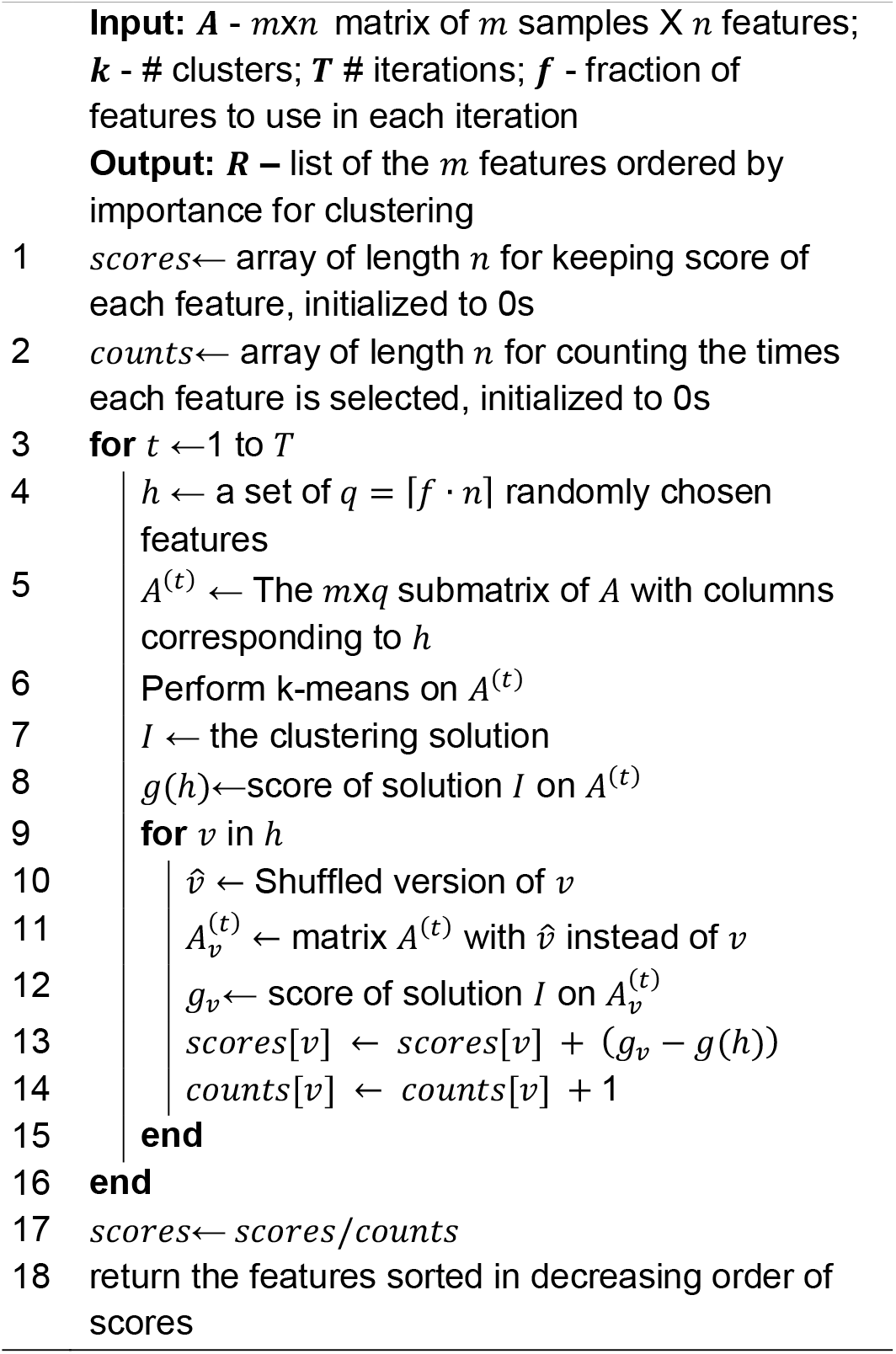

#### Time complexity

For simplicity we analyze FRIGATE for continuous features. The runtime of k-means is *O*(*mqkc*) for *m* samples, *q* features, *k* clusters and up to *c* iterations. We sample in each iteration *q* features. For each k-means run we perform *i* initializations. Therefore, the runtime of the k-means executions in each FRIGATE iteration is *O*(*mqkci*). Other than k-means runs, in each iteration, for each of the *q* features, we shuffle the values of its features over the full cohort in *O*(*m*) and recalculate the solution score *d*_*v*_ in *O*(*m*). The overall runtime of an iteration is *O*(*mqkci* + *mq*) = *O*(*mqkci*). We perform up to *T* iterations, so the overall runtime is *O*(*mTqkci*). As *q* = *f n* and *f* is a constant, we can write the runtime as *O*(*mTnkci*).

##### Algorithm 2

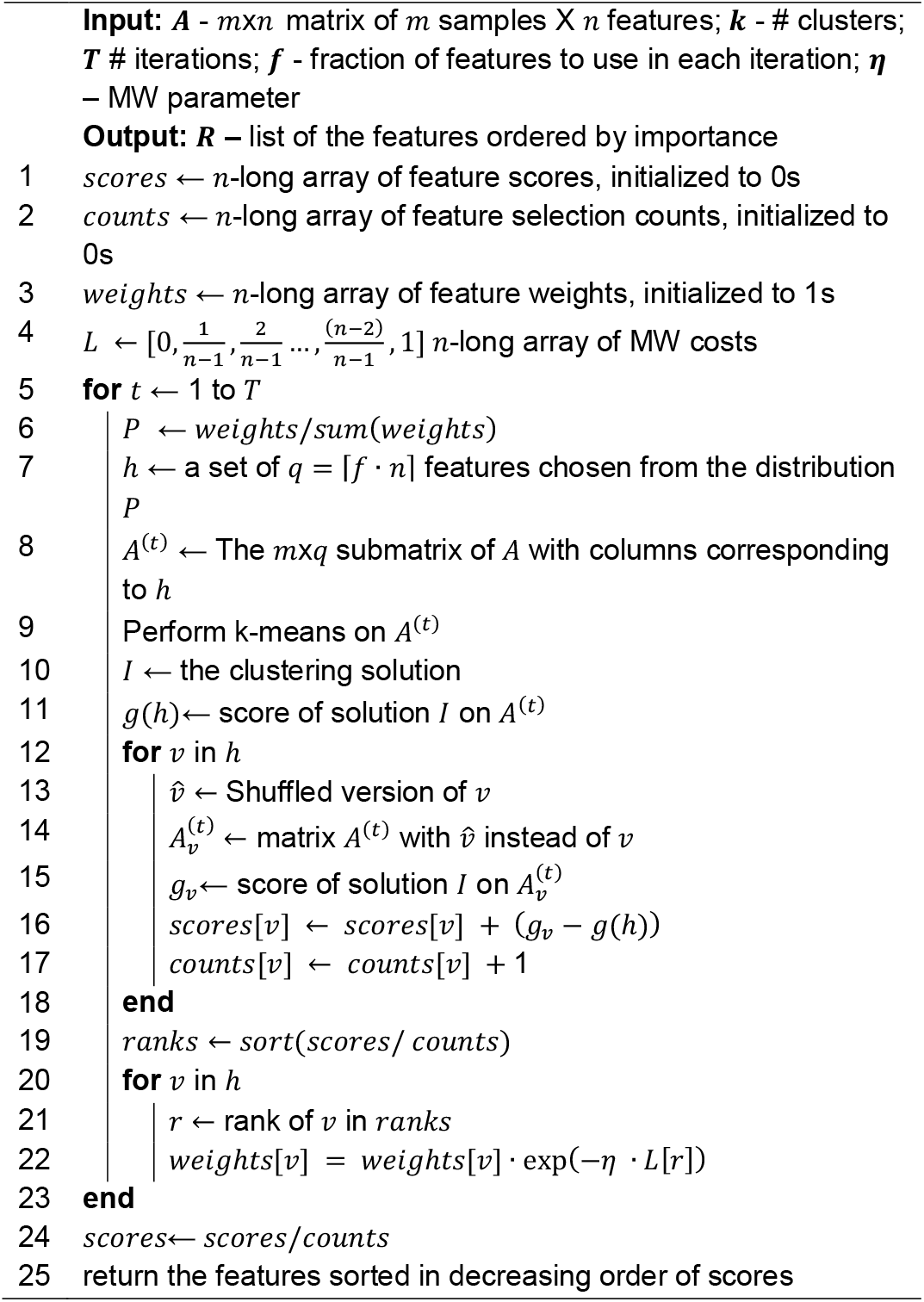

The FRIGATE-MW Algorithm

### 3.2 The Frigate-MW algorithm

MW offers a potentially smarter way to choose the features in FRIGATE for each clustering subproblem instead of choosing them randomly. Algorithm 2 (Algorithm 2) shows the version of FRIGATE that uses MW for continuous features, which we call FRIGATE-MW. We define an n-long array *L* so that *L*(*i*) = (*i* − 1)/(*n* − 1). At each iteration we rank the features by their scores so far and use the ranks and *L* to determine 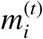 (Equation 4). If the rank of feature *i* at iteration *t* is *r* then 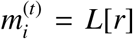. The weights of features that were not selected in the iteration remain unchanged. For the next iteration the sum of weights at the *t*-th iteration. we select features from distribution 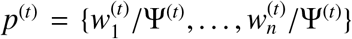 where 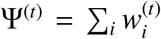 is the sum of weights at the *t*-th iteration.

#### Time complexity

In each iteration we update the weights of the *q* participating features in constant time for each feature and sort the array of weights in *O*(*n* log(*n*)). The overhead of MW for each iteration is thus *O*(*n* log(*n*) + *q*) = *O*(*n* log(*n*)), since *q* < *n*. The total runtime of each iteration in FRIGATE-MW is: *O*(*mqkci* + *n* log(*n*)). Therefore, the total runtime is: *O*(*mTqkci* + *Tn* log(*n*)) = *O*(*Tn*(*mkci* + log(*n*))). Altogether, the increase in the runtime over FRIGATE is modest. However, in FRIGATE-MW the iterations cannot be programmed to run in parallel, in contrast to FRIGATE. For both versions of the algorithm we used *T* = 2*n* and *f* = 0.1, and for FRIGATE-MW we used η = 0.5. See full analyses of the parameter choice in Supplementary 1. Note that k-means is sensitive to outliers, which can impact the results of FRIGATE. Users should consider how to address outliers before running FRIAGTE.

### 3.3 Simulation

We performed simulations in order to test the algorithms in situations where the true clustering and the informative features are known. The simulations were along the same lines of [14].The parameters of the simulation are:

- *k* – number of clusters
- *c* – number of samples in each cluster
- *α* – number of informative features
- *β* – number of non-informative features
- µ– distribution parameter
- σ – correlation coefficient between features

#### Simulating continuous data

For each cluster *j* ∈ {0, …, *k* − 1}, we construct *c* vectors of length *n* = *α* + *β* from multivariate normal distribution, where *α* features are sampled from a normal distribution with mean of *j*µ. The other *β* features are sampled from a normal distribution with mean 0 for all clusters and therefore represent the non-informative features. Thus, the mean vector of a sample in the *j*-th cluster is: µ _*j*_ = [(*j*µ)_(*α*×1)_, 0_(*β*×1)_].

Next, we define a covariance matrix, parameterized by σ, used to create correlations between the different features. The covariance matrix Σ is identical for all clusters:

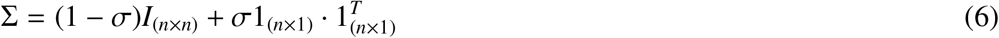

The *n* × (*kc*) data matrix *A* then undergoes z-score normalization for each feature. This step is needed when working in the medical domain as the values of different features can be of different magnitude.

For **simulating mixed data** we add three more parameters:

- *α*_*categorical*_ – number of informative categorical features
- *β*_*categorical*_– number of non-informative categorical features
- *p* – probability of choosing the right categorys

We assume *l* categorical features labeled 0, 1, …, *l* − 1. For the informative features of a sample in the *j*-th cluster, we choose the value (category) *j* with probability *p* and a value from {0, …, *l* − 1}\{*j*}with probability 1 − *p* where the value is chosen uniformly at random. For the non-informative features we choose a random value uniformly from {0, …, *l* − 1}. The simulation of the continuous features is done as described above, and we concatenate the two matrices into a single input matrix. In our simulation we used *p* = 0.95.

### 3.4 Evaluation Measures

On real datasets, often the true feature ranking is unknown, even when a true clustering is known. We developed a method to evaluate a ranking when the clustering is known. It is inspired by [14, 12, 13] and uses the Adjusted Rand Index (ARI), an established measure for comparing two clustering solutions [24]. ARI has range of [−1, 1] with a higher value for better match of the solutions. The reasoning behind this is that a better ranking is anticipated to yield a clustering solution that is more similar to the correct one when employing solely the top-ranked features. This is because it ranks the most informative features at the top.

We run k-means on the subset of the data containing only the *j* top ranked features. The clustering produced is compared to the true labels using ARI. For ranking algorithm *a*, denote the resulting ARI by Γ(*a, j*). The process is repeated for *j* = 1, …, *n*. This gives a value for each *j* (see Figure 1below). We developed two scores that summarize the values across all *j*.

**Figure 1.**
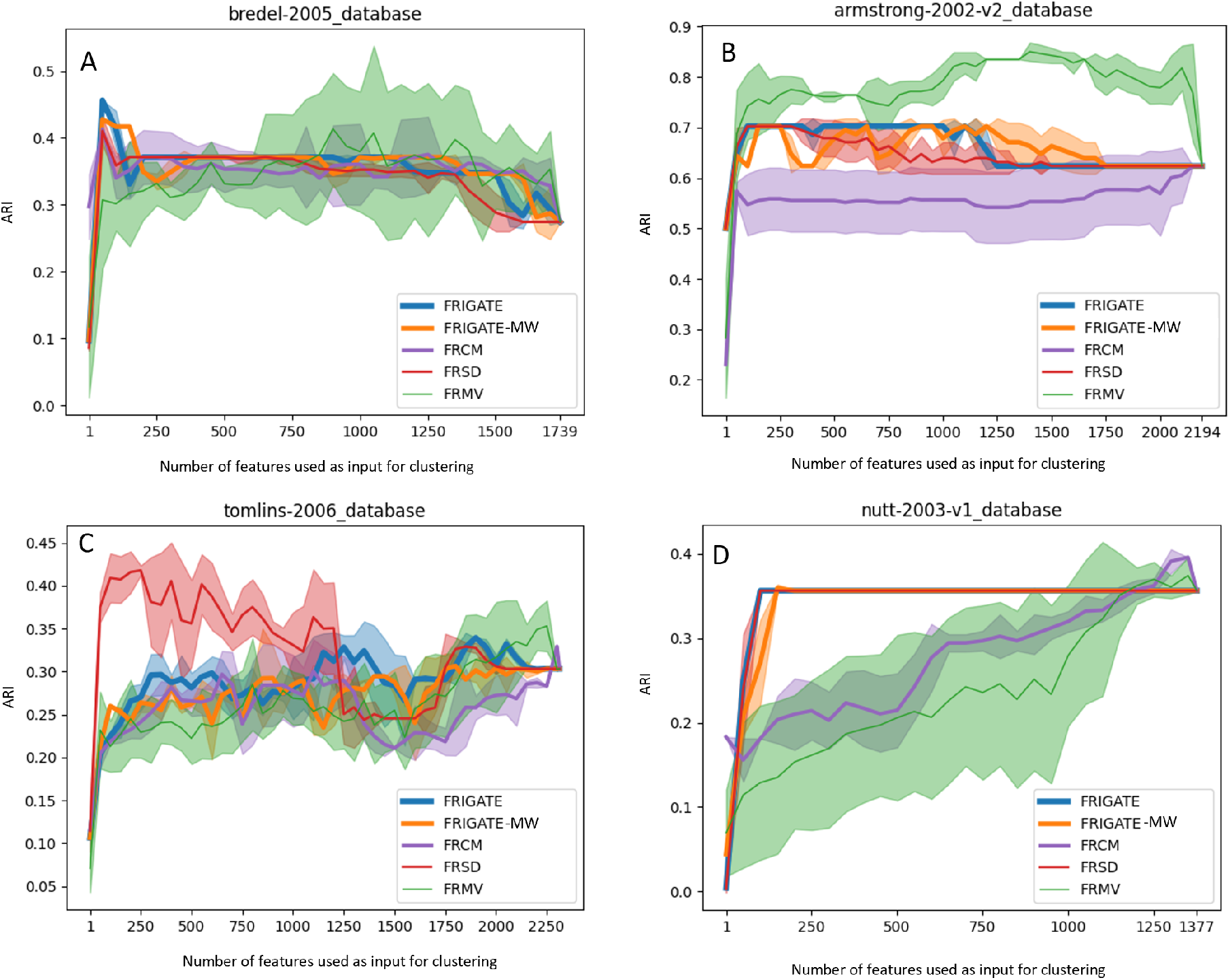
A-D: Performance on datasets 1-4 respectively. For every algorithm, for each *j*, the feature ranking produced by the algorithm was used to cluster the data using the top *j* features, and the resulting clusters were compared to the known ones using ARI. X axis: *j*. Y axis: ARI. Results are average of ten runs; the light-colored sleeve shows ±1 std for each plot.

The first score compares the performance of *M* different algorithms *a*_1_, …, *a*_*M*_ on the same dataset. For each *j*, we rank the algorithms based on their scores Γ(*a*_*i*_, *j*), from 1 for the top performer to *M*. Denote by *rank*(*a, j*) the rank of algorithm *a* on the top *j* features. For simplicity of the description, we assume there are no ties. The *weighted rank* (*WR*) of algorithm *a* is defined as:

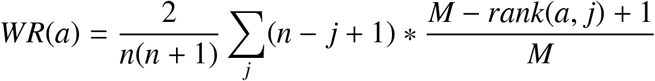

Hence, the second factor in the sum ranges from 1 for the top ranked algorithm to 1/*M* for the worst ranked, and the first factor gives higher weights to smaller *j*-s, from *n* for the first feature to 1 for the last ranked. The factor 2/(*n*(*n* + 1)) rescales the total sum to [0, 1].

The second score evaluates a single algorithm. Let *ARI*(*a, j*) be the ARI of algorithm *a* using the top *j* features. The *weighted ARI* (*WARI*) of the algorithm is defined as:

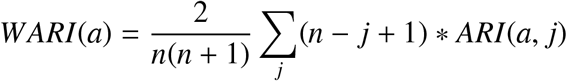

Again, the second factor in the sum is higher for better match, and the first factor in the sum gives higher weights for higher ranked features. Both scores can be generalized to handle ties and also situations where not all values of *j* are tested, e.g., when there are too many features.

## 4. Results

We measured the performance of FRMV, FRSD, FRCM, FRIGATE and FRIGATE-MW on simulated and real data. Since no implementations were provided for the extant algorithms, we implemented them as described in [14, 12, 13]. The code for all five algorithms is available in:https://github.com/Shamir-Lab/FRIGATE. We used k-means as implemented in [26]for continuous data with 100 k-means++ initializations in each run, For mixed data we used the k-prototype implementation in [27]. For FRMV we used the linear correlation as the relevance measure. For FRSD we used silhouette as implemented in Scikit learn [26]. The number of clusters *k* in FRIGATE and for FRMV was chosen with the elbow method implemented as suggested in [28].For a full analysis of the effect of *k* see Supplementary 7.

### Simulated data

We simulated data as described in Section 3with 200 samples and 100 features of which 20 are informative. The scenarios tested were two or four equal-sized clusters, µ = 0.5 and 1, and σ = 0, 0.05, 0.2, and 0.5. See Supplementary 3 for analysis of the choice of σ. We ran the algorithms on data with and without z-score normalization. The *recognition rate* is defined as the fraction of informative features in the top 20 ranked features. Results for *k* = 4 and varying µ and σ are shown in Table 1. In all cases, the elbow method chose *k* = 2. On normalized data FRCM performed best and FRIGATE-MW second. On non-normalized data FRIGATE-MW was best. FRMV scored poorly in all cases. FRSD scored poorly in most normalized scenarios, but high in most non-normalized scenarios. In general, smaller values of µ account for harder cases, and normalized data are more challenging than non-normalized data. Larger values of *k* were easier (results not shown). For FRIGATE, FRIGATE-MW and FRCM high levels of correlation (σ ≥ 0.2) cause a drop in performance. FRSD and to some extant

**Table 1:** Recognition rates for simulated data generated with *k* = 4, *c* = 50, *α* = 20, *β* = 80.

| parameters | $\mu = 0.5$<br>$\sigma = 0$ | $\mu = 0.5$<br>$\sigma = 0.05$ | $\mu = 0.5$<br>$\sigma = 0.2$ | $\mu = 0.5$<br>$\sigma = 0.5$ | $\mu = 1$<br>$\sigma = 0$ | $\mu = 1$<br>$\sigma = 0.05$ | $\mu = 1$<br>$\sigma = 0.2$ | $\mu = 1$<br>$\sigma = 0.5$ |
| --- | --- | --- | --- | --- | --- | --- | --- | --- |
| normalized |  |  |  |  |  |  |  |  |
| FRIGATE | 0.98±0.03 | 0.91±0.07 | 0.46±0.17 | 0.09±0.08 | 1±0 | 1±0 | 0.97±0.05 | 0.09±0.08 |
| FRIGATE-MW | 1±0 | 1±0 | 0.62±0.33 | 0.19±0.15 | 1±0 | 1±0 | 1±0 | 0.01±0.02 |
| FRCM | 1±0 | 1±0 | 0.72±0.15 | 0.35±0.18 | 1±0 | 1±0 | 1±0 | 0.99±0.03 |
| FRSD | 0.06±0.04 | 0.06±0.04 | 0.11±0.06 | 0.38±0.17 | 0.01±0.02 | 0±0 | 0.04±0.05 | 0.32±0.08 |
| FRMV | 0.13±0.16 | 0.13±0.13 | 0.25±0.16 | 0.16±0.12 | 0.05±0.1 | 0.03±0.03 | 0.09±0.12 | 0.06±0.16 |
| non-normalized |  |  |  |  |  |  |  |  |
| FRIGATE | 0.99±0.02 | 0.98±0.03 | 0.76±0.12 | 0.7±0.15 | 1±0 | 1±0 | 1±0 | 1±0 |
| FRIGATE-MW | 1±0 | 1±0.02 | 0.94±0.08 | 0.31±0.14 | 1±0 | 1±0 | 1±0 | 1±0 |
| FRCM | 1±0 | 0.99±0.02 | 0.82±0.1 | 0.45±0.2 | 1±0 | 1±0 | 1±0 | 1±0 |
| FRSD | 0.79±0.06 | 0.74±0.11 | 0.77±0.07 | 0.89±0.06 | 1±0.02 | 1±0 | 1±0 | 1±0 |
| FRMV | 0.2±0.19 | 0.12±0.2 | 0.23±0.2 | 0.11±0.08 | 0.02±0.06 | 0.02±0.04 | 0.13±0.2 | 0.1±0.15 |

FRMV show opposite behavior, where results improve under extreme correlation levels. The performance of FRSD and FRMV varies more than that of FRIGATE and FRCM across different scenarios (see Supplementary 4).

As 20% of the features were informative, a recognition rate below 0.2 reflects performance that is worse than random ordering of features. FRMV often showed such results, FRSD scored low for most of the normalized cases with low correlation levels, and FRIGATE scored below random levels in the extreme correlation setting. Although correlation levels of σ ≥ 0.2 are unrealistic for medical data, future work should be directed to understand these results. FRCM rarely dropped significantly below random levels (results not shown).

### Real data

We tested the five algorithms on eleven real genomic and EMR datasets from different sources for which a known or natural clustering was available. The datasets are described in Table 2.The genomic Datasets 1-4 were used in a benchmark of clustering [35].Of the eight EMR datasets, Datasets 5-7 are from the MIMIC-III repository [3], 8 from Zigong Fourth People’s Hospital [33],and 9-11 from the eICU repository [34].All were downloaded from PhysioNet [36]. The input features used were continuous, containing lab tests (“labs”), age and length of stay in the hospital (days in MIMIC, minutes in eICU) and Apache score in eICU. (For the specific set of features that were used in each dataset, please see the cited papers.) For each lab, we included only the patient’s first measurement during the ICU stay. For each patient we included data from a single ICU stay. For the MIMIC datasets ICD-9 diagnosis codes were used for labeling the patients. For the eICU datasets, diagnoses and Apache score parameters were used as categorical variables and for labeling. Labs that were missing in >70% of the cohort were removed. To remove potential outliers, we z-scored each continuous measurement across the cohort, and removed patients who had any lab with |*z* − *score*| ≥ 3. We then applied the Iterative Imputer from [26]inspired by the MICE algorithm [37]on the raw data to complete missing values and performed z-score normalization. The cohorts and clusters that we constructed were:

**Table 2:** Real datasets benchmarked. *k* - # clusters. () in column 4: cluster sizes; in column 5: # continuous and categorical features. G: genomic; E: EMR; C: continuous; M: mixed.

| # | Dataset and source | k | # samples | # features | Type |
| --- | --- | --- | --- | --- | --- |
| 1 | Bredel-2005 [29] | 3 | 50 (31, 14, 5) | 1739 | G, C |
| 2 | Armstrong-2002-v2 [30] | 3 | 72 (24, 20, 28) | 2194 | G, C |
| 3 | Tomlins-2006 [31] | 5 | 50 (27, 20, 32, 13, 12) | 2315 | G, C |
| 4 | Nutt-2003-v1 [32] | 4 | 50 (14, 7, 14, 15) | 1377 | G, C |
| 5 | Young cancer patients [3] | 2 | 161 (122, 39) | 70 | E,C |
| 6 | Young healthy patients [3] | 2 | 110 (84, 26) | 47 | E,C |
| 7 | Newborns [3] | 2 | 5286 (1534, 3752) | 29 | E,C |
| 8 | Heart failure [33] | 2 | 169 (68, 101) | 77 | E,C |
| 9 | Intubated patients [34] | 2 | 441 (136, 305) | 157 (87, 70) | E,M |
| 10 | Short stay at ICU [34] | 2 | 570 (487, 83) | 79 (59, 20) | E,M |
| 11 | Young patients [34] | 2 | 232 (138, 94) | 86 (72, 14) | E,M |

- Dataset 5 – patients aged 18-40 with ICD-9 cancer diagnosis, divided into two clusters by length of stay: 122 patients who were discharged alive after <18 days in ICU, and 39 patients who either died during the ICU stay or were discharged after ≥18 days.
- Dataset 6 – “healthy” patients: individuals aged 20-30 with no ICD-9 codes of cancer, benign tumors, hypertension, cardiac disease, endocrine related disease, or hepatitis who stayed up to one day at ICU. They were divided into two clusters by sex: 84 males and 26 females.
- Dataset 7 – newborns in two clusters: 1534 with jaundice and 3752 without jaundice.
- Dataset 8 - heart failure patients from two age groups: 68 patients aged 29-49 and 101 patients of ages 89-100.
- Dataset 9 – intubated patients aged ≥70 divided according to status at discharge of “Alive”, 305 patients, and “Expired”, 136 patients.
- Dataset 10 – patients who stayed <1 day in ICU, separated by age groups: 487 patients aged 18 to 80, and 83 patients aged 80+.
- Dataset 11 – patients aged 18-30 separated by length of stay: 138 who stayed >4.5 days or expired, and 94 who stayed ≤4.5 days and were discharged alive.

Table 3 shows the performance of the five tested algorithms in terms the WR and WARI scores on datasets 1-11. We ran each algorithm on each dataset ten times and computed mean and standard deviation of the WARI and WR scores. Overall, FRIGATE and FRIGATE-MW show best performance, followed by FRCM. FRMV and FRSD performed worse than the other three, although FRSD performed best on two datasets and FRMV on one. The results using the two different scores are consistent on most datasets. FRIGATE shows a slight advantage over FRIGATE-MW, although the latter was best performer in some cases. The WARI measure tended to distinguish better than WR between the best performer and the lesser ones, as WARI uses the ARI values only and not their relative ranks.

**Table 3:**
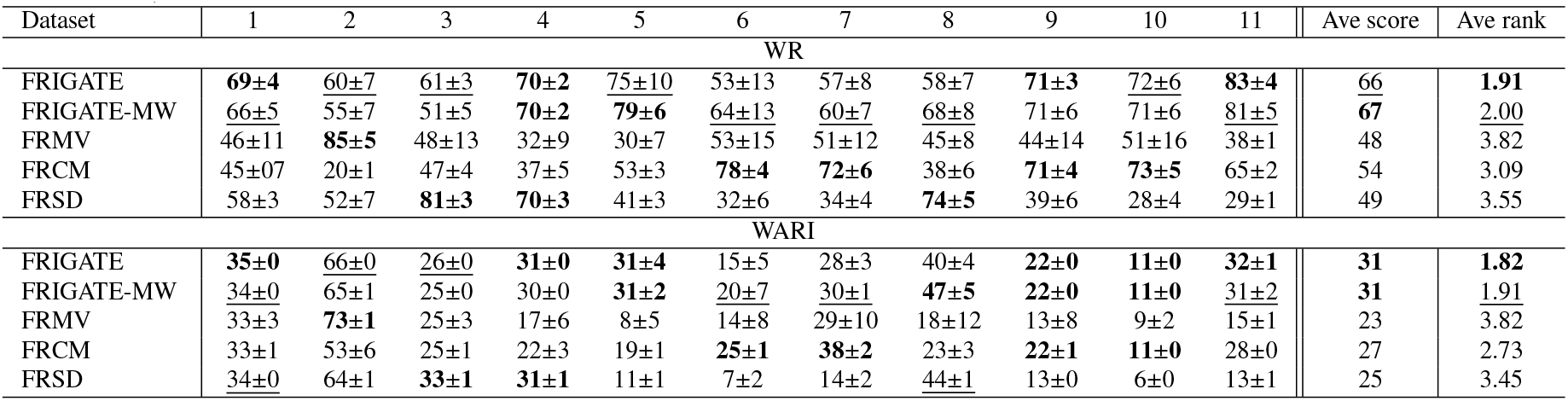
Performance of the algorithms on datasets 1-11, as measured by WR and WARI scores. Results are means and std of ten runs. Numbers are multiplied by 100 for easier visualization. Bold: top performer for the dataset, underlined: second best.

Detailed results for the genomic Datasets 1-4 are shown in Figure 1. These datasets have thousands of features and only a few dozen samples each. Thus it is expected that many features do not carry information relevant to the clustering. Indeed, for most algorithms and datasets we see a sharp rise in the ARI from 1 to about 100 features and little improvement for adding more features. In all cases the value chosen by the elbow method for FRIGATE and FRMV was k=2. FRIGATE, FRIGATE-MW and FRSD had comparable and generally good performance, reaching maximum ARI of 0.35-0.7 in most cases. FRSD performed markedly better than the other methods on Dataset 3. FRCM performed worse in most cases, with slow gradual increase in ARI. FRMV performed best on Dataset 2, with wide variance across repetitions.

Detailed results for EMR Datasets 5-8 are shown in Figure 2. Again we see a sharp rise in ARI for the top ranked 20-30% of the features in most cases. By and large, FRMV and FRSD showed slower increase, and FRCM had mixed behavior, with slow increase in Datasets 5 and 8 but the fastest in 6 and 7. FRIGATE and FRIGATE-MW performed consistently well except for Dataset 7, which has very few features. For Dataset 8, FRSD performed comparably to FRIGATE and FRIGATE-MW and even better in some thresholds (for more details see Supplementary 5).

**Figure 2.**
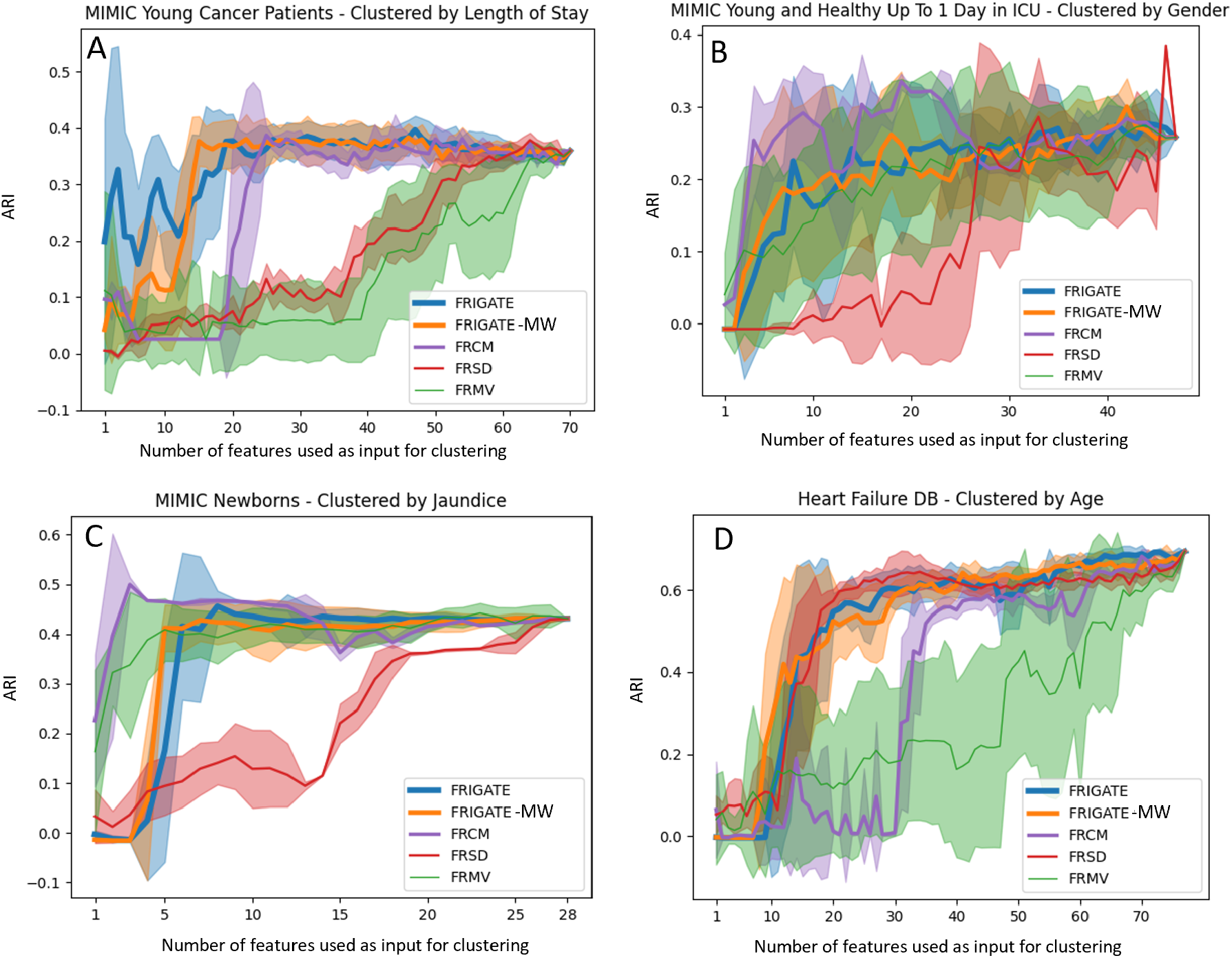
A-D: Performance on datasets 5-8 respectively. See Figure 1captions for details.

The results for EMR Datasets 9-11 using the continuous features are shown in Figure 3A,C,E.The same trends are observed: FRIGATE, FRIGATE-MW and FRCM perform best, FRMV has a large variance, and FRSD performs poorly.

**Figure 3.**
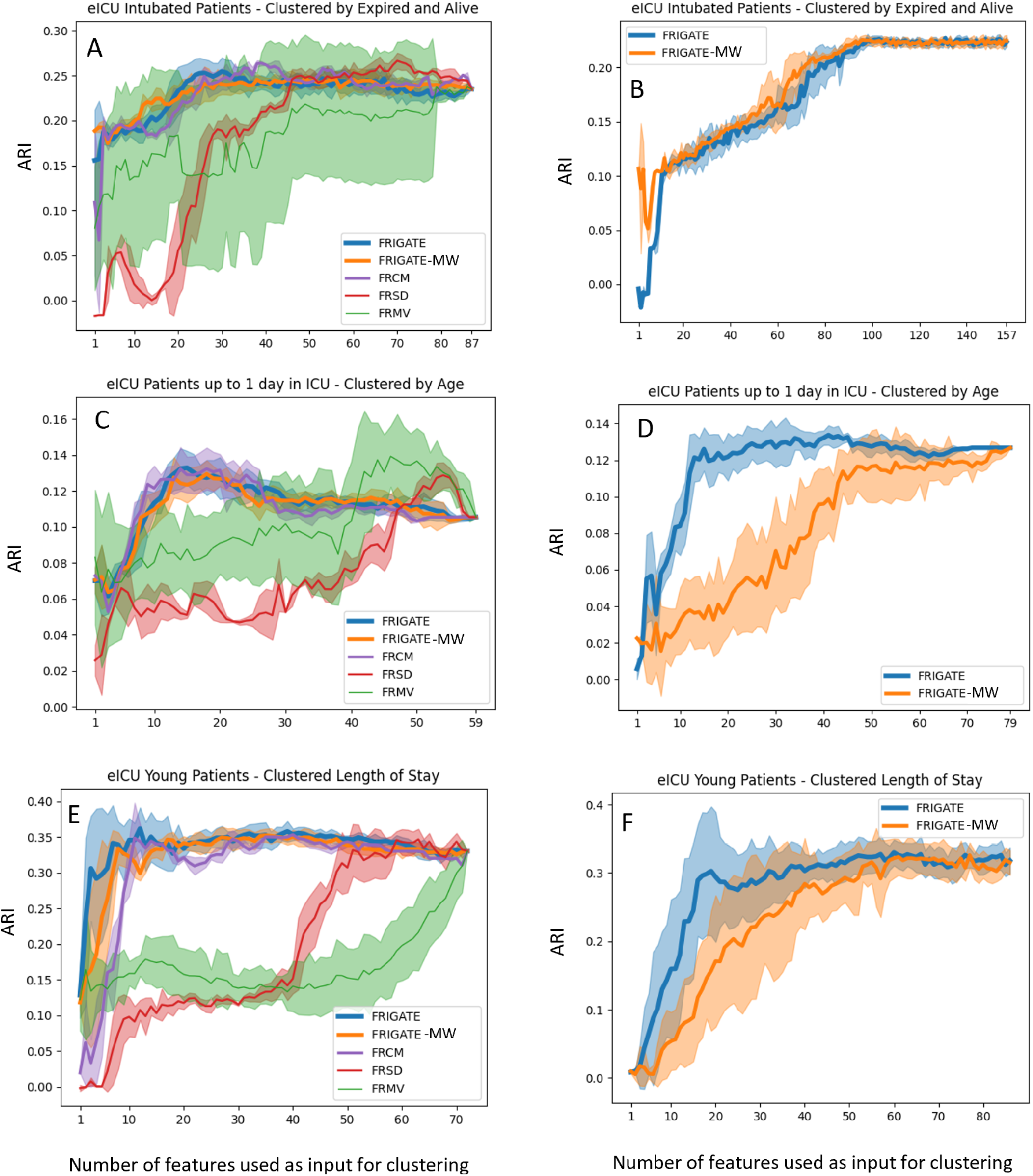
A,C,E: Performance on datasets 9,10,11 respectively, using the continuous features. B,D,F: Performance of FRIGATE and FRIGATE-MW using also the categorical features. See Figure 1captions for details.

Using also categorical features could be tested only for FRIGATE and FRIGATE-MW. To choose the best γ factor in Equation eq:dxymixed, we tested different values of γ and looked for a change in the ARI using the full set of features in comparison to using only the continuous features. A change in ARI means a different composition of the clusters caused by the categorical features. Based on this analysis (Supplementary 1.3) we chose γ = 6. The results on the mixed data (Figure 3B,D,F)did not show much improvement, and in some cases worsened.

### Clinical evaluation

For an independent evaluation of the ranking’s clinical relevance, we chose to focus on Dataset 6, since a recent study reported sex-based differences in lab tests [38]. We focused on twelve features that were also available in Dataset 6 and according to [38]fulfil:

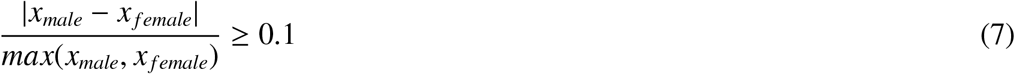

where *x*_*i*_ is the mean value of feature *x* for sex *i*. We call these the top features. A hypergeometric test between the 12 first ranked features according to FRIGATE and the top features obtained a p-value of 0.136. The same test for FRIGATE-MW gave p=0.034. We also calculated the minimum hypergeometric score (mHG), as used in [39],for calculating the significance without determining in advance the threshold for the test and accounting for multiple testing. For FRIGATE the mHG was obtained for 13 features, with p=0.07. For FRIGATE-MW the threshold was 10 features with p=0.01. Hence, according to this test, only FRIGATE-MW’s ranking was clinically significant. See full results in Supplementary 6. Note however that test is limited by the very small number of top features and by possible differences between the cohorts of [38]and of Dataset 6.

### Runtime

Table 4shows the runtimes of the tested algorithms on Datasets 1-8. Results are means of multiple runs. Standard deviations were 1-3% of the mean and are omitted for visibility. FRIGATE and FRIGATE-MW were slower on Datasets 1-4, which have many features and a few samples, but fast on the EMR datasets, which have less features. To speed up these algorithms when the number of features is very large, one can decrease the number of iterations and increase the number of features per iteration *f*, as we observed that a larger *f* improves the ranking (results not shown). FRCM ran fast on Datasets 1-4, but its runtime increased sharply on Dataset 7, which has many samples. This algorithm has a set number of iterations, and produces an *m* × *m* matrix for each feature, which is both time and space consuming. Note also the slowdown of FRSD on Dataset 7. FRMV was fastest in all but one dataset.

**Table 4:**
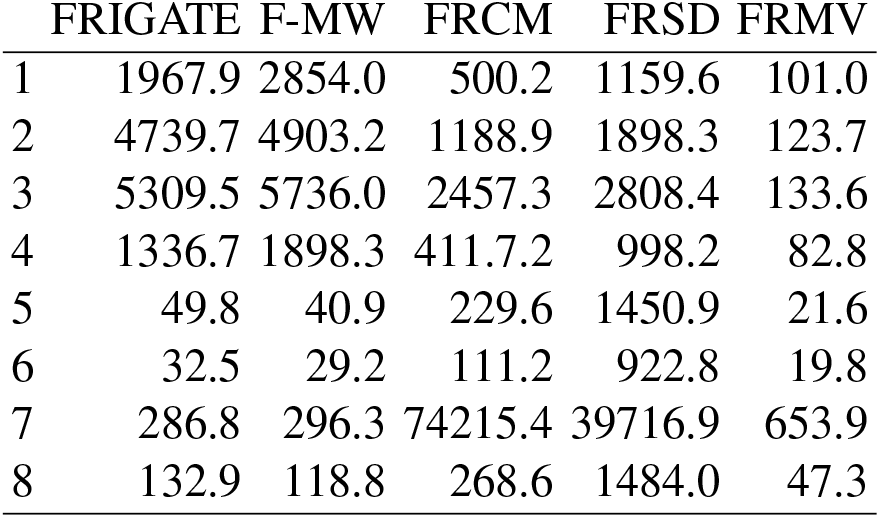
Runtime in seconds for Datasets 1-8.

## 5. Discussion

We presented FRIGATE, a new feature ranking algorithm for clustering, aimed for medical data, and FRIGATE-MW, a variant of FRIGATE that uses multiplicative weights for feature weighting. To the best of our knowledge, this is the first use of game-theoretic approaches for the problem. We also proposed two new global scores for EFR algorithms, offering consolidated summary scores that enhance the evaluation process compared to previous approaches that assess the top *j* features separately for each *j*.

We compared FRIGATE and FRIGATE-MW to three extant EFR methods: FRMV, FRCM and FRSD, on simulated and on real EMR datasets. FRIGATE and FRIGATE-MW performed robustly well, and had short runtime unless the number of features was extremely large. On real datasets, FRIGATE performed best and FRIGATE-MW was a close runner-up, followed by FRCM. We make available software implementations of all five algorithms. The ranking of features in medical data allows physicians to interpret the clustering results clinically and to assign better meaning to patient subgroups that the clustering reveals.

The simulation results presented interesting behaviors of the tested algorithms. FRSD and FRMV seem to improve, while FRIGATE and FRCM performed worse with higher correlation levels. Intuitively, it should be harder to set apart the informative features from the full set of features when high correlation levels are present. Our hypothesis is that enforcing extreme levels of correlation between all features shaped the data so that the differences between features are better captured by the changes in the silhouette score, which is incorporated in FRSD. This necessitates further study.

FRIGATE and FRIGATE-MW had different behavior on simulated and real data. On simulated data, the two algorithms performed comparably, with a mild advantage to FRIGATE-MW. On real data FRIGATE performed slightly better. Future work should consider using alternative MW cost functions.

FRIGATE and FRIGATE-MW can handle mixed data, with both continuous and categorical features. They can also analyze pure categorical data, by using the k-modes algorithm for clustering [21].In our tests, adding the categorical features did not improve the results, and in some cases even harmed them. Alternative ways to include categorical data should be investigated.

The number of clusters *k* is needed as input in FRIGATE and FRIGATE-MW. However, on simulated data, the results were stable even when the input value of *k* was much larger than the real *k* (Supplementary 7). Future research should aim to remove the required input *k*. Note that for single disease EMR data we expect a small value of *k* (corresponding to the number of disease subtypes) so the full range of relevant *k* values can be tested. In every FRIGATE iteration the values of a each participating feature are shuffled once in order to eliminate its contribution to the clustering solution. Multiple shuffles can give a better estimate of the feature’s contribution, at the cost of higher runtime. This tradeoff can be studied further.

Our study has several limitations. The comparison to the extant EFR algorithms was based on our implementation, since code of these algorithms was not available. A limitation in the evaluation of EMR data was the validity of the clusters that we produced.

Heterogeneous cohorts like these of MIMIC and eICU contain multiple overlapping patient subgroups, which may confound clustering attempts and their evaluation. Also, all the EMR datasets that we analyzed contained two clusters. More analysis is needed on medical datasets with a larger number of clusters. Moreover, in order to measure performance more confidently, one needs a gold standard EMR data with both known clustering and a known feature ranking. Finally, a test of the algorithms on real EMR data where both the number of patients and the number of features is very large, and sparsity of features is modest, is desired.

## 6. Conclusion

FRIGATE is a novel ensemble algorithm that performs feature ranking for clustering, outperforming extant methods. The development of FRIGATE was primarily based on medical data, in order to face the special characteristics of these data and provide a tool for future researchers, mostly in uncovering new disease sub-types.

## Supporting information

all supplementary files

## Data Availability

All real data used in this paper are from publicly available sources. See Table 2 for details.

## 7. CRediT authorship contribution statement

**Eran Shpigelman**: Conceptualization, Methodology, Software, Validation, Formal analysis, Data Curation, Writing - Original Draft, Writing - Review Editing. **Ron Shamir**: Conceptualization, Methodology, Validation, Formal analysis, Writing - Original Draft, Writing - Review Editing, Supervision, Funding acquisition.

