## Supplementary material for "A feature ranking algorithm for clustering medical data": all supplementary files

### 1. Supplementary 1 – Parameter choice

#### 1.1 Parameter choice for FRIGATE-MW

FRIGATE-MW has three parameters: $\boldsymbol{T}$, the number of iterations, $\boldsymbol{f}$, the fraction of the features used in each iteration, and $\boldsymbol{\eta}$, the MW parameter. If the input is mixed data there is also $\boldsymbol{\gamma}$, the K-prototypes parameter. We seek values of $T$ and $f$ that are small in order to increase the efficiency of the algorithm, but still produce good results.

For testing the performance of FRIGATE-MW with different hyper-parameters, we built a simulation as described in chapter 3.3, with the following parameters: $k = 4$ clusters of size $c = 50$ each, $\alpha = 20$ informative features and $\beta = 80$ non-informative ones. The statistical parameters were $\mu = 1$ and $\sigma = 0.05$. The value of $\sigma$ was chosen to produce similar correlations to those observed in real data (see details in Supplementary 4).

Figure S1 shows the average accurate recognition rate results of 10 simulations for different values of $f$, $T$ and $\eta$. As expected, more iterations and larger $f$ produce better results. We can see that for 200 iterations the results are perfect for all values of $\eta$ with $f \geq 0.1$, and the results for 100 iterations are a close second (perfect results for $\eta$=0.25 and 0.5, and 0.995 for 1). Increasing $\eta$ from 0.25 to 1 improves the results. For the same parameters of the simulation but $\mu = 4$ (which causes the clusters to be more distinct) we observed similar results (Figure S2).

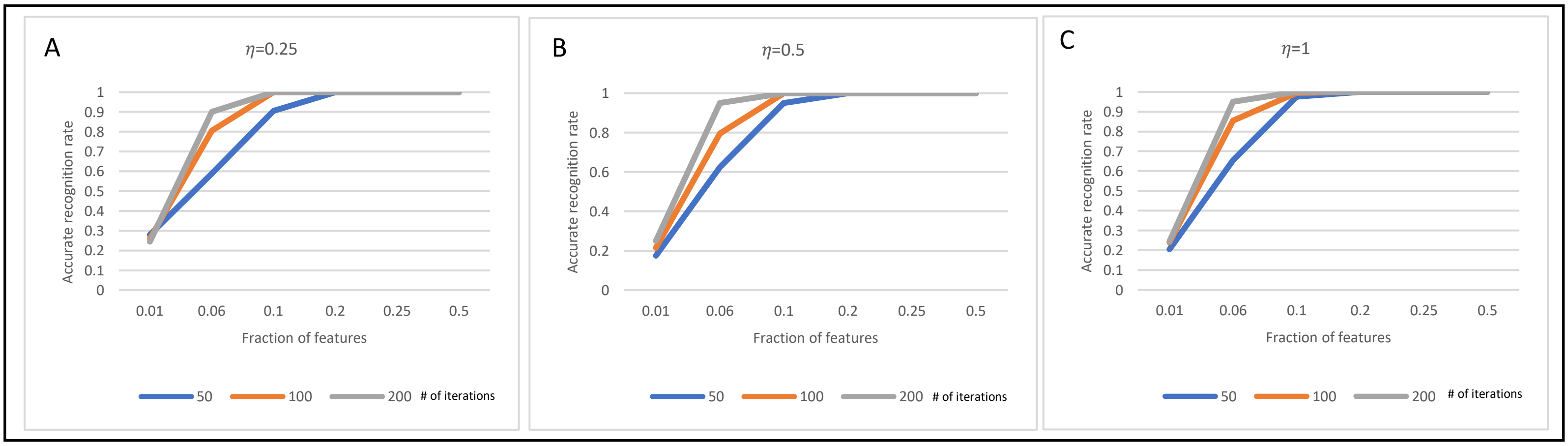

Figure S1. Performance of FRIGATE-MW on simulated data for different hyper-parameters. The simulation had 4 clusters, 50 samples in each cluster, and 100 features of which 20 were informative. The differences between the clusters were set by $\mu = 1, \sigma = 0.05$. The graphs show the accurate recognition rate of the algorithm, averaged over 10 runs. Plot color: number of iterations. X axis: fractions of features participating in each iteration. A, B and C present results for values of $\eta = 0.25$, 0.5 and 1 respectively.

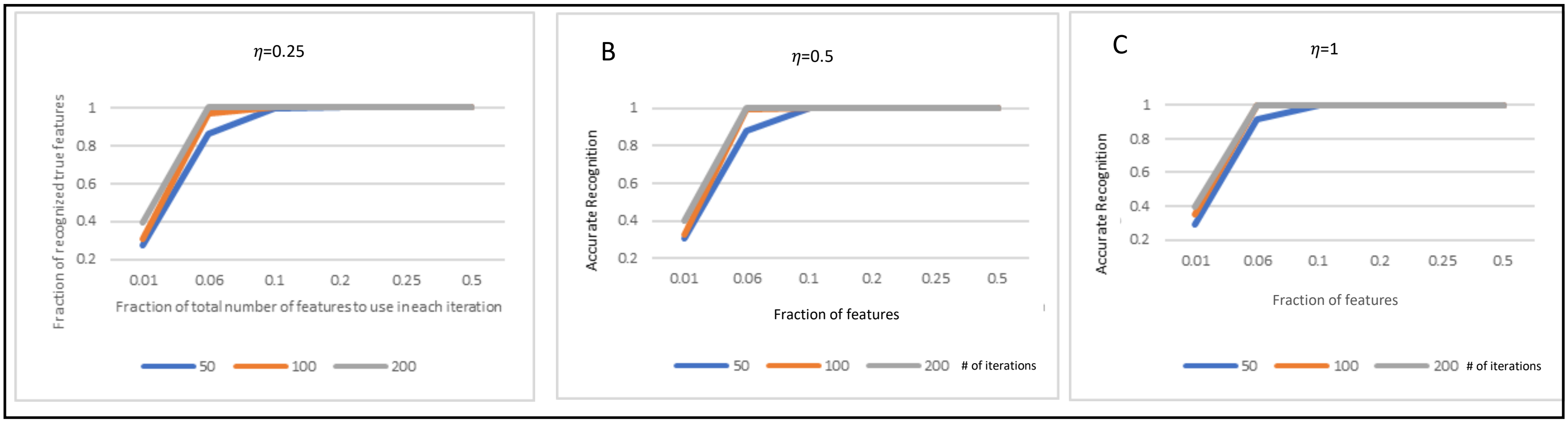

Figure S2. Performance of FRIGATE-MW with different hyper-parameters. The simulation setup and presented results are as in Figure S1, but here we used $\mu = 4$.

We also tested the scenario of two clusters with 100 samples in each cluster ($k = 2, c = 100$) and kept the rest of the parameters unchanged (Figure S3). The trends are similar to Figure S1, but we can see that the overall scores are slightly worse. Still, the combination with $f = 0.2, \eta \geq 0.5$ and $T = 200$ achieved perfect results. A similar simulation with $\mu = 4$ achived high results in all scenarios (Figure S4). In another simulation that used values similar to [6]: $k = 4,\ c = 50\ and\ n = 950$ the results were perfect for all parameters tested. We therefore chose to fix the hyper-parameters to $f = 0.1$, and $T = 2 \cdot |V|$.

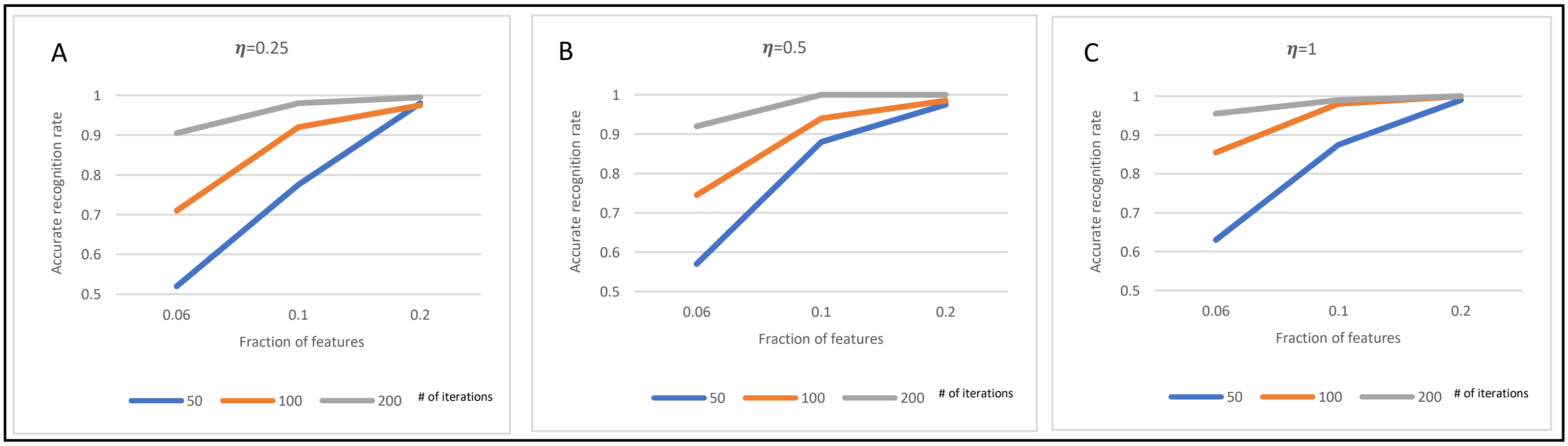

Figure S3. Performance of FRIGATE-MW on simulated data for different hyper-parameters. The simulation setup and presented results are as in Figure S1, but here there were 2 clusters and 100 samples in each cluster.

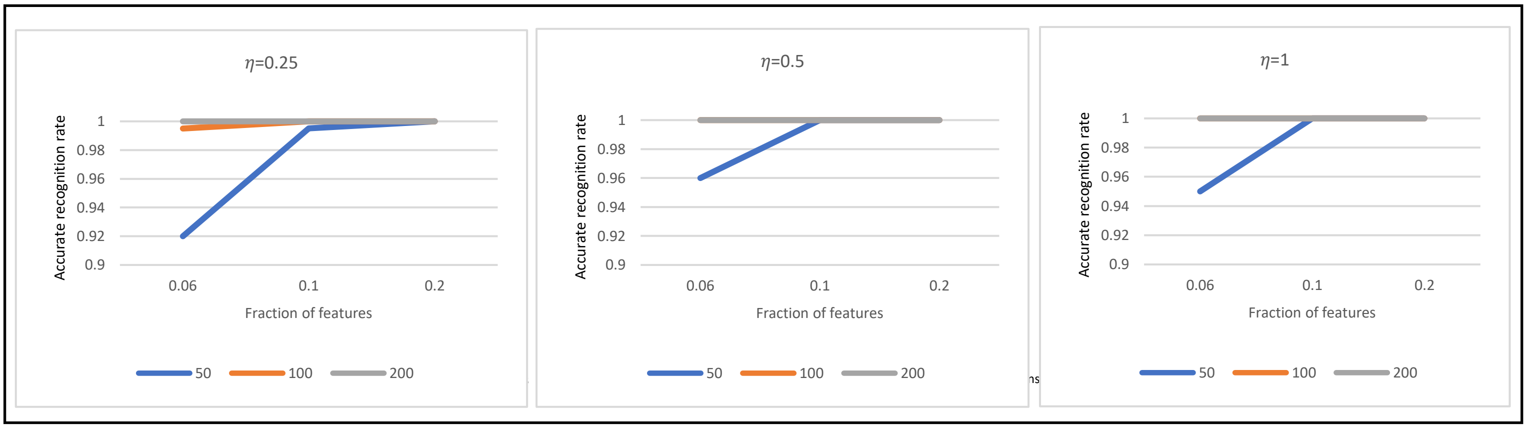

Figure S4. Performance of FRIGATE-MW on simulated data for different hyper-parameters. The simulation setup and presented results are as in Figure S3, but here we used $\mu = 4$..

#### 1.2 Parameter choice for FRIGATE

The results of FRIAGTE in the same simulations are shown in Figure S5 for $k = 2, 4$ and $\mu = 1{,}4$. We can see that the algorithm without MW improves more slowly with the number of iterations and is in general inferior. This can be seen especially in the hardest case of $\mu = 1; k = 2$. Based on these results we chose as default values $f = 0.1, T = 2 \cdot |V|$ for the non-MW algorithm as well.

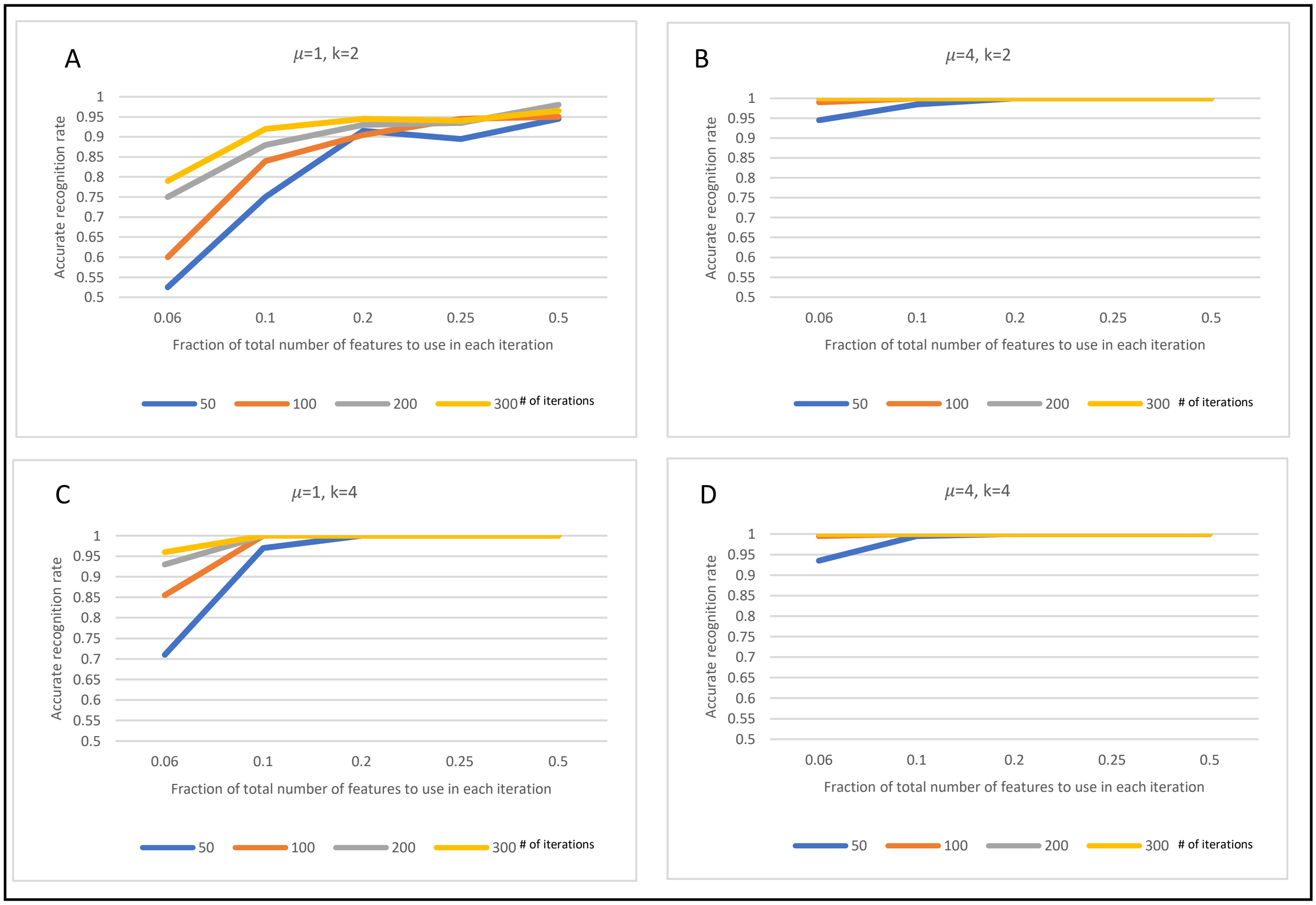

Figure S5. Performance of FRIGATE on simulated data for different hyper-parameters. Simulation parameters: $\sigma$=0.05, total number of samples=200, 100 feature of which 20 are informative. A: $\mu = 1, k = 2$. B: $\mu = 4, k = 2$. C: $\mu = 1, k = 4$. D: $\mu = 4, k = 4$.

#### 1.3 Weight parameter $\gamma$ for categorical features

Next, we wanted to determine the value of the weight parameter $\gamma$ in the k-prototypes algorithm, when using mixed data. We performed simulations with 100 continuous features of which 20 are informative ($\alpha = 20, \beta = 80$), $\sigma = 0.05$ and 200 samples equally divided between clusters. The other parameters were tested in multiple options: $k = \{2,4\}$ (accounted for $c = 100$ and $c = 50$ respectively), $\mu = \{1,4\}$, {100 categorical features of them 20 informative ($\alpha_{categorical} = 20, \beta_{categorical} = 80$), 50 categorical features of them 10 informative ($\alpha_{categorical} = 10,\ \beta_{categorical} = 40$)}. We ran each scenario 10 times and for each type of features calculated the accurate recognition rate in the top thirty or forty features, depending on the total number of informative features.

The average results of FRIGATE and FRIGATE-MW for $\mu = 1$ and $k = 2$ are shown in Tables S2-S5. We can see that in all cases $\gamma = 1$ performed perfectly and $\gamma = 2$ near perfectly in both scenarios, unrelated to the proportion of continuous and categorical features. As expected, extreme values of $\gamma$, which favor one type of features over the other, were detrimental. For $k = 4$ and $\mu = 4$ the results were nearly perfect in all scenarios and thus not informative (results not shown).

| Table S2-FRIGATE-MW | $\gamma$ | | | |
|---|---|---|---|---|
| | **0.5** | **1** | **2** | **4** |
| Continuous Features | 1 | 1 | 0.99 | 0.845 |
| Categorical Features | 1 | 1 | 1 | 1 |

| Table S3-FRIGATE-WW | $\gamma$ | | | |
|---|---|---|---|---|
| | **0.5** | **1** | **2** | **4** |
| Continuous Features | 1 | 1 | 0.975 | 0.38 |
| Categorical Features | 0.82 | 1 | 1 | 1 |

| Table S4-FRIGATE | $\gamma$ | | | |
|---|---|---|---|---|
| | **0.5** | **1** | **2** | **4** |
| Continuous Features | 1 | 1 | 0.96 | 0.595 |
| Categorical Features | 0.99 | 1 | 1 | 1 |

| Table S5-FRIGATE | $\gamma$ | | | |
|---|---|---|---|---|
| | **0.5** | **1** | **2** | **4** |
| Continuous Features | 1 | 1 | 1 | 0.93 |
| Categorical Features | 0.97 | 1 | 1 | 1 |

Tables S2-S5. Average accurate recognition rates in 10 simulation runs, for mixed data by FRIGATE and FRIGATE-MW for different values of the weight parameter $\gamma$ in the k-prototypes algorithm. Simulation parameters: $\sigma = 0.05$, $c_j$ = 100, $\alpha = 20$, $\beta = 80$, $\mu = 1$, $k = 2$. Tables 3,5: $\alpha_{categorical} = 20$, $\beta_{categorical} = 80$. Tables 4,6: $\alpha_{categorical} = 10$, $\beta_{categorical} = 40$.

The results of this simulation suggest that $\gamma = 1$ is the favorable value. However, this may not hold for all cases. We can see that for $\gamma > 2$ the results are poor, regardless of the proportion between continuous and categorical features, in contradiction with pervious publications [7]. More research is needed regarding the choice of $\gamma$.

### 2. Supplementary 2 – Demonstration of a FRIGATE

For better understanding of the FRIGATE process, we demonstrate it graphically. We simulated data as described in chapter 3.3, with two continuous features, two clusters ($k = 2$), and 100 samples in each cluster, and simulation parameters $\mu = 4$, $\sigma = 0$. Figure S6 shows the data, where each axis is a feature and the samples are colored by cluster membership. We simulated three scenarios:

1. Both features are informative for the clustering solution (Figure S6A).
2. Only one feature is informative (Figure S6B)
3. Both features are not informative (Figure S6C).

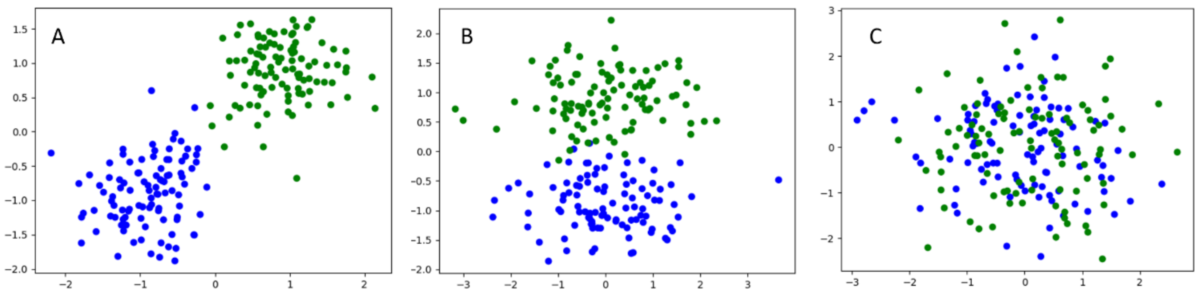

D

| Scenario | Solution Score | Scores after shuffling values of F1 | | Scores after shuffling values of F2 | |
|---|---|---|---|---|---|
| | | Solution | Feature | Solution | Feature |
| A | 81.76 | 395.24 | 313.48 | 368.09 | 286.33 |
| B | 237.28 | 237.79 | 0.51 | 541.33 | 304.05 |
| C | 256.48 | 374.07 | 114.59 | 407.04 | 150.56 |

Figure S6. Illustrations of simulations with two clusters. In the simulation, there are 100 samples in each cluster, $\mu = 4$, $\sigma = 0$, and two features. A-C: features are represented by the axes. Each color represents a different cluster. A: both features are informative for clustering, B: only the feature represented by the $y$ axis is informative, C: both features are not informative. D: Demonstration of FRIGATE iteration on the data of A-C. Solution score refers to line 8 in Algorithm 1, feature's score refers to line 13 in Algorithm 1. F1 is represented by the x-axis in Figure1 A-C and F2 is represented by the y-axis. We can see that the informative features received higher scores than the non-informative ones.

Next, we performed an iteration of the FRIGATE algorithm, using the centroids obtained from the clustering solution on the two features, to show the differences in scores in each scenario (Figure S6D):

1. When the two features were informative, the solution score (line 8 in Algorithm 1) was 81.76, and the scores of the features (line 13 in Algorithm 1) were 313.48 and 286.33. Both feature

scores are high, and the difference can result from the randomness in shuffling the values (line 10 in Algorithm 1) or from the simulation that might have produced one feature that is more informative than the other.

2. When only one feature was informative, the solution score was 237.28, and the feature scores were 0.51 for the non-informative features and 304.05 for the informative feature.
3. When the two features were non-informative, the solution score was 256.48, and the feature scores were 117.59 and 150.57.

In Figures S7-9 we demonstrate graphically an iteration in each scenario. In all scenarios the informative features scored much higher than the non-informative ones. Notice that the differences in scores are due to the initial solution score of each scenario – the poor results of scenario 3 already produced a relatively high solution score, so the ability of any feature to score high is limited.

<u>Scenario 1 – two informative features</u>

Figure S7A shows the results of k-means clustering of the simulated data with two informative features (line 6 in Algorithm 1), with a solution score of 81.76. Figure S7B shows the data after shuffling the values of the x-coordinate feature (line 10 in Algorithm 1). The new solution score is 395.24 (line 12 in Algorithm 1) and a feature score of 313.48. Figure S7C shows the data after shuffling the values of the y-coordinate feature, which led to a new solution score of 368.09 and a feature score of 286.33.

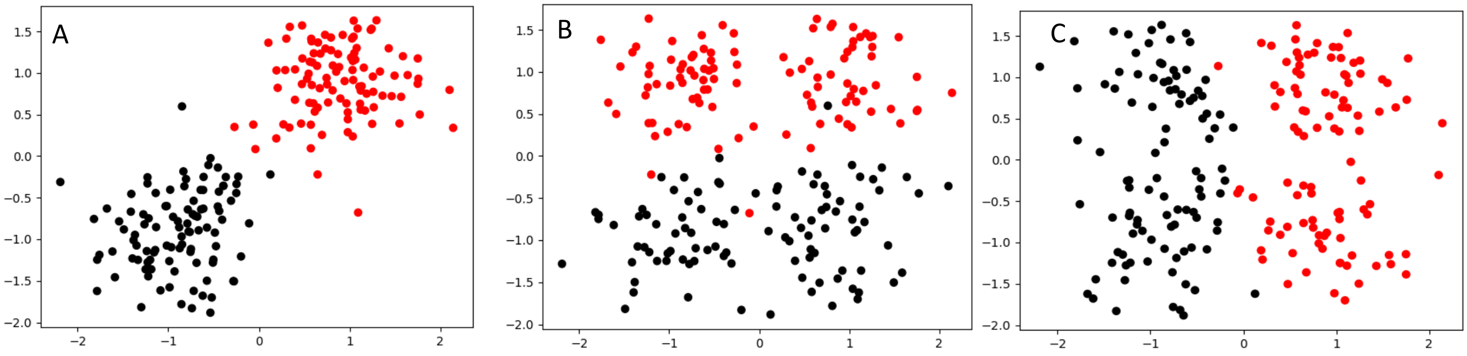

Figure S7. Illustration of the different steps of FRIGATE for scenario 1 of two informative features. A – A clustering solution of the data (line 6 in Algorithm 1) colored by clusters labels. The solution score is 81.76 (line 8 in Algorithm 1). B- Results of shuffling the x-coordinate (lines 10-13 in Algorithm 1). The solution score increased dramatically to 286.33. C- Results of shuffling the y-coordinate feature. We see again a great change from S1A with a solution score of 313.48.

<u>Scenario 2 – one informative feature and one non-informative feature</u>

Figure S8A shows the results of k-means clustering of the data (line 6 in Algorithm 1), with a solution score of 237.28. Figure S8B shows the data after shuffling the values of the non-informative feature (line

10 in Algorithm 1). The shuffled data has an almost identical solution score of 237.79 (line 12 in Algorithm 1) and a feature score of 0.51. Figure S8C shows the sample locations after shuffling the values of the informative feature, which gives a new solution score of 541.33 and a feature score of 304.05.

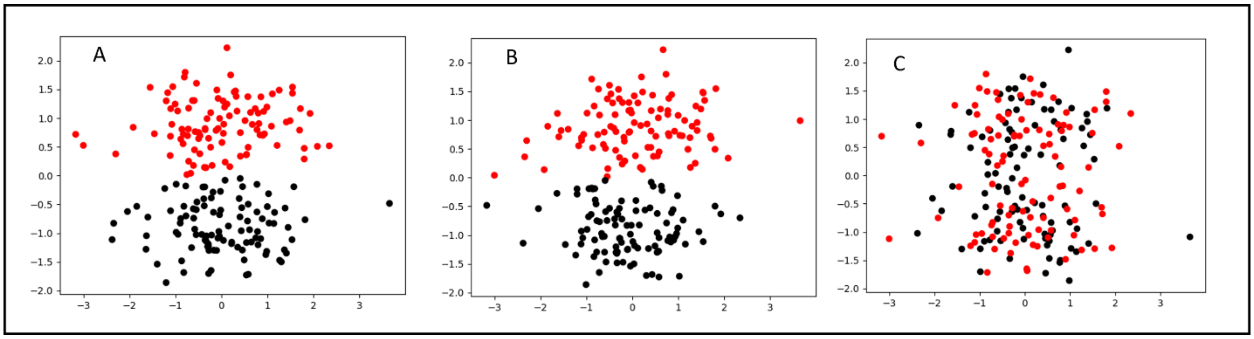

Figure S8. Illustrations of the different steps of FRIGATE for scenario 2, where one feature is informative (y axis) and one is non-informative (x axis). A – a clustering solution of the data (line 6 in Algorithm 1) colored by clusters labels. The solution score is 237.28 (line 8 in Algorithm 1). B- Results of shuffling the x coordinates, representing the non-informative feature (lines 10-13 in Algorithm 1). The solution score is similar to A and the feature's score is 0.51. C- Results of shuffling the y coordinates, representing the informative feature. An increase in the solution score led to a feature's score of 304.5.

<u>Scenario 3 – two non-informative features</u>

Figure S9A shows the results of k-means clustering of the data where both features are non-informative. The solution score is 256.48. Figure S9B shows the data after shuffling the values of y-coordinate feature. The new solution score is 374.07 (line 12 in Algorithm 1) and a feature score of 114.59. Figure S9C shows the data after shuffling the values of the x-coordinate feature, which led to a solution score of 407.04 and feature score of 150.56.

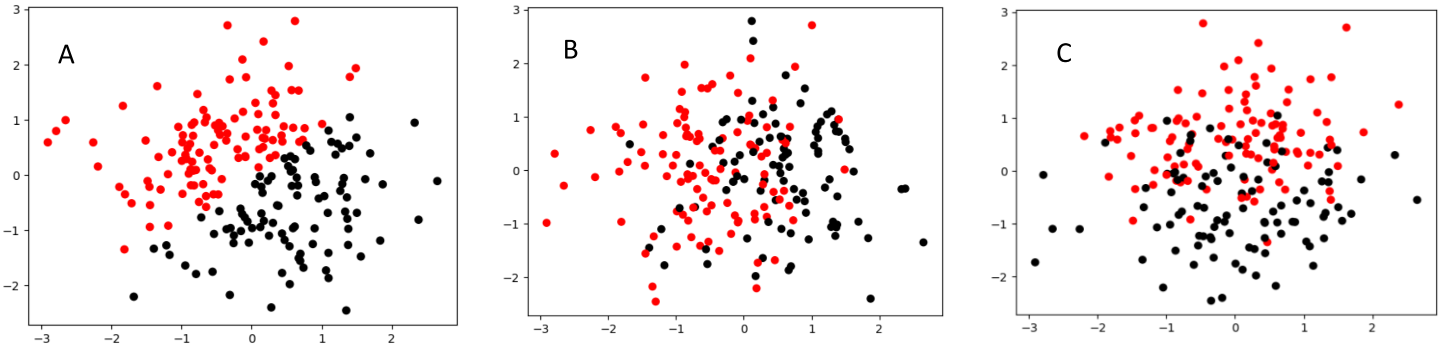

Figure S9. Illustration of the different steps of FRIGATE for scenario 3 of two non-informative features. A – A clustering solution of the data (line 6 in Algorithm 1) colored by clusters labels. The solution score is 256.48 (line 8 in Algorithm 1). B- Results of shuffling the x-coordinate feature (lines 10-13 in Algorithm 1). The solution score changed to 150.56. C- Results of shuffling the y-coordinate feature. The solution score is similar to Figure S2B, 114.59.

### 3. Supplementary 3 - Choosing the value of $\sigma$

We tested the average Pearson correlation between the different features when constructing the simulation using different values of $\sigma$. We tested three scenarios and the average results of 10 runs for $\mu = 2$ are presented in Figure S10:

- Scenario 1 - 200 samples divided into 4 clusters ($k = 4$) with 20 informative features and 80 non-informative.
- Scenario 2 – same as scenario 1 with 2 clusters ($k = 2$).
- Scenario 3 – the simulation suggested in [6]: 200 samples, 4 clusters ($k = 4$), 50 informative features, 950 non informative features.

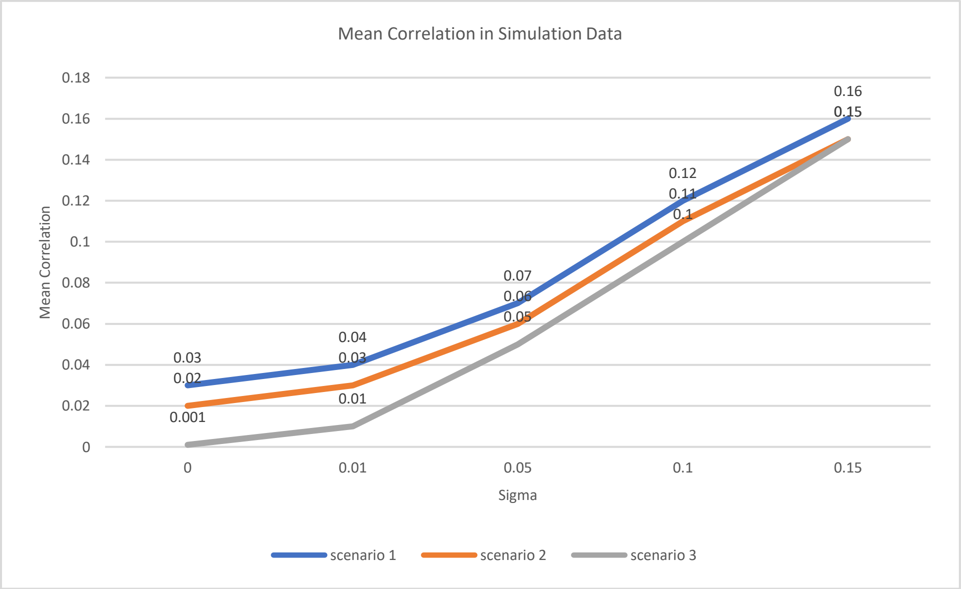

Figure S10. Testing the average Pearson correlation (X axis) in the data in different simulation scenarios, with different values of $\sigma$ (Y axis). Description of the scenarios is found above.

For $\mu = \{1,4\}$ we received nearly identical results (not shown).

Next, we tested the average Pearson correlation in real data sets. The four genomic cancer datasets as used in [12] and presented in the Results (chapter 4, Datasets 1-4) have an average Pearson correlation of 0.07 with STD of 0.03. The value of $\sigma$ that achieved similar correlation in the data is 0.05, so other than the simple case of $\sigma = 0$ we desided to test also $\sigma = 0.05$, and $\sigma = 0.2$ as a case of extreme correlation. We also tested the highly unrealistic $\sigma = 0.5$ since it was used in [6].

### 4. Supplementary 4 – simulation results

Simulated data with 200 samples and 100 features of which 20 are informative, divided into two or four equal-sized clusters, and tested $\mu = \{0.5,1,2,4\}$ and $\sigma = \{0,0.05,0.2,0.5\}$, with and without z-score normalization. Results of the accurate recognition rate, supplement to Table 1.

| $k = 4$, normalized | $\mu = 2$ $\sigma = 0$ | $\mu = 2$ $\sigma = 0.05$ | $\mu = 2$ $\sigma = 0.2$ | $\mu = 2$ $\sigma = 0.5$ | $\mu = 4$ $\sigma = 0$ | $\mu = 4$ $\sigma = 0.05$ | $\mu = 4$ $\sigma = 0.2$ | $\mu = 4$ $\sigma = 0.5$ |
|---|---|---|---|---|---|---|---|---|
| FRIGATE | $1 \pm 0$ | $1 \pm 0$ | $1 \pm 0.02$ | $0.06 \pm 0.1$ | $1 \pm 0$ | $1 \pm 0$ | $1 \pm 0$ | $0.1 \pm 0.12$ |
| FRIGATE-MW | $1 \pm 0$ | $1 \pm 0$ | $1 \pm 0$ | $0 \pm 0$ | $1 \pm 0$ | $1 \pm 0$ | $1 \pm 0$ | $0.3 \pm 0.46$ |
| FRCM | $1 \pm 0$ | $1 \pm 0$ | $1 \pm 0$ | $1 \pm 0$ | $1 \pm 0$ | $1 \pm 0$ | $1 \pm 0$ | $1 \pm 0$ |
| FRSD | $0 \pm 0$ | $0 \pm 0$ | $0.14 \pm 0.1$ | $0.66 \pm 0.12$ | $0.05 \pm 0.06$ | $0.05 \pm 0.04$ | $0.25 \pm 0.1$ | $0.66 \pm 0.11$ |
| FRMV | $0 \pm 0$ | $0 \pm 0$ | $0.04 \pm 0.11$ | $0.43 \pm 0.3$ | $0 \pm 0$ | $0 \pm 0$ | $0 \pm 0$ | $0.24 \pm 0.31$ |

Table 6. Performance on simulated data, with $k = 4$ and z-score normalized data.

| $k = 4$, Non-normalized | $\mu = 2$ $\sigma = 0$ | $\mu = 2$ $\sigma = 0.05$ | $\mu = 2$ $\sigma = 0.2$ | $\mu = 2$ $\sigma = 0.5$ | $\mu = 4$ $\sigma = 0$ | $\mu = 4$ $\sigma = 0.05$ | $\mu = 4$ $\sigma = 0.2$ | $\mu = 4$ $\sigma = 0.5$ |
|---|---|---|---|---|---|---|---|---|
| FRIGATE | $1 \pm 0$ | $1 \pm 0$ | $1 \pm 0$ | $1 \pm 0$ | $1 \pm 0$ | $1 \pm 0$ | $1 \pm 0$ | $1 \pm 0$ |
| FRIGATE-MW | $1 \pm 0$ | $1 \pm 0$ | $1 \pm 0$ | $1 \pm 0$ | $1 \pm 0$ | $1 \pm 0$ | $1 \pm 0$ | $1 \pm 0$ |
| FRCM | $1 \pm 0$ | $1 \pm 0$ | $1 \pm 0$ | $1 \pm 0$ | $1 \pm 0$ | $1 \pm 0$ | $1 \pm 0$ | $1 \pm 0$ |
| FRSD | $1 \pm 0$ | $1 \pm 0.02$ | $1 \pm 0$ | $1 \pm 0$ | $1 \pm 0$ | $1 \pm 0$ | $1 \pm 0$ | $1 \pm 0$ |
| FRMV | $0 \pm 0$ | $0 \pm 0$ | $0 \pm 0$ | $0 \pm 0$ | $0 \pm 0$ | $0 \pm 0$ | $0 \pm 0$ | $0 \pm 0$ |

Table 7. Performance on simulated data, with $k = 4$ and non-normalized data.

| $k = 2$, normalized | $\mu = 0.5$ $\sigma = 0$ | $\mu = 0.5$ $\sigma = 0.05$ | $\mu = 0.5$ $\sigma = 0.2$ | $\mu = 0.5$ $\sigma = 0.5$ | $\mu = 1$ $\sigma = 0$ | $\mu = 1$ $\sigma = 0.05$ | $\mu = 1$ $\sigma = 0.2$ | $\mu = 1$ $\sigma = 0.5$ |
|---|---|---|---|---|---|---|---|---|
| FRIGATE | $0.34 \pm 0.09$ | $0.39 \pm 0.11$ | $0.29 \pm 0.09$ | $0.15 \pm 0.07$ | $0.96 \pm 0.04$ | $0.89 \pm 0.06$ | $0.46 \pm 0.11$ | $0.16 \pm 0.11$ |
| FRIGATE-MW | $0.38 \pm 0.11$ | $0.37 \pm 0.18$ | $0.22 \pm 0.1$ | $0.14 \pm 0.1$ | $1 \pm 0.02$ | $1 \pm 0$ | $0.56 \pm 0.22$ | $0.15 \pm 0.2$ |
| FRCM | $0.49 \pm 0.08$ | $0.42 \pm 0.11$ | $0.23 \pm 0.08$ | $0.14 \pm 0.08$ | $1 \pm 0.02$ | $0.98 \pm 0.02$ | $0.66 \pm 0.11$ | $0.29 \pm 0.16$ |
| FRSD | $0.17 \pm 0.07$ | $0.15 \pm 0.05$ | $0.23 \pm 0.08$ | $0.26 \pm 0.08$ | $0.09 \pm 0.05$ | $0.12 \pm 0.07$ | $0.15 \pm 0.08$ | $0.37 \pm 0.1$ |
| FRMV | $0.19 \pm 0.11$ | $0.19 \pm 0.07$ | $0.18 \pm 0.07$ | $0.22 \pm 0.12$ | $0.1 \pm 0.12$ | $0.14 \pm 0.13$ | $0.12 \pm 0.09$ | $0.26 \pm 0.19$ |

Table 8. Performance on simulated data, with $k = 2$ and z-score normalized data.

| $k = 2$, normalized | $\mu = 2$ $\sigma = 0$ | $\mu = 2$ $\sigma = 0.05$ | $\mu = 2$ $\sigma = 0.2$ | $\mu = 2$ $\sigma = 0.5$ | $\mu = 4$ $\sigma = 0$ | $\mu = 4$ $\sigma = 0.05$ | $\mu = 4$ $\sigma = 0.2$ | $\mu = 4$ $\sigma = 0.5$ |
|---|---|---|---|---|---|---|---|---|
| FRIGATE | $1 \pm 0$ | $1 \pm 0$ | $0.94 \pm 0.07$ | $0.11 \pm 0.14$ | $1 \pm 0$ | $1 \pm 0$ | $1 \pm 0$ | $0.21 \pm 0.17$ |
| FRIGATE-MW | $1 \pm 0$ | $1 \pm 0$ | $1 \pm 0$ | $0.24 \pm 0.39$ | $1 \pm 0$ | $1 \pm 0$ | $1 \pm 0$ | $0.1 \pm 0.3$ |
| FRCM | $1 \pm 0$ | $1 \pm 0$ | $1 \pm 0$ | $1 \pm 0$ | $1 \pm 0$ | $1 \pm 0$ | $1 \pm 0$ | $1 \pm 0$ |
| FRSD | $0 \pm 0$ | $0 \pm 0$ | $0.07 \pm 0.07$ | $0.55 \pm 0.1$ | $0.26 \pm 0.07$ | $0.28 \pm 0.07$ | $0.35 \pm 0.11$ | $0.37 \pm 0.08$ |
| FRMV | $0.25 \pm 0.27$ | $0.19 \pm 0.25$ | $0.06 \pm 0.15$ | $0.17 \pm 0.16$ | $0.21 \pm 0.27$ | $0.09 \pm 0.15$ | $0.01 \pm 0.02$ | $0.24 \pm 0.32$ |

Table 9. Performance on simulated data, with $k = 2$ and z-score normalized data.

| $k = 2$, Non-normalized | $\mu = 0.5$ $\sigma = 0$ | $\mu = 0.5$ $\sigma = 0.05$ | $\mu = 0.5$ $\sigma = 0.2$ | $\mu = 0.5$ $\sigma = 0.5$ | $\mu = 1$ $\sigma = 0$ | $\mu = 1$ $\sigma = 0.05$ | $\mu = 1$ $\sigma = 0.2$ | $\mu = 1$ $\sigma = 0.5$ |
|---|---|---|---|---|---|---|---|---|
| **FRIGATE** | $0.41 \pm 0.1$ | $0.37 \pm 0.1$ | $0.27 \pm 0.11$ | $0.3 \pm 0.12$ | $0.96 \pm 0.04$ | $0.93 \pm 0.04$ | $0.82 \pm 0.09$ | $0.54 \pm 0.14$ |
| **FRIGATE-MW** | $0.45 \pm 0.12$ | $0.43 \pm 0.13$ | $0.33 \pm 0.11$ | $0.32 \pm 0.11$ | $0.99 \pm 0.02$ | $1 \pm 0.02$ | $0.89 \pm 0.18$ | $0.79 \pm 0.13$ |
| **FRCM** | $0.51 \pm 0.15$ | $0.38 \pm 0.11$ | $0.25 \pm 0.07$ | $0.19 \pm 0.06$ | $1 \pm 0$ | $0.99 \pm 0.02$ | $0.75 \pm 0.14$ | $0.42 \pm 0.1$ |
| **FRSD** | $0.32 \pm 0.06$ | $0.36 \pm 0.08$ | $0.32 \pm 0.1$ | $0.37 \pm 0.1$ | $0.74 \pm 0.08$ | $0.72 \pm 0.09$ | $0.73 \pm 0.04$ | $0.8 \pm 0.07$ |
| **FRMV** | $0.18 \pm 0.09$ | $0.22 \pm 0.12$ | $0.21 \pm 0.09$ | $0.2 \pm 0.09$ | $0.14 \pm 0.17$ | $0.18 \pm 0.25$ | $0.28 \pm 0.2$ | $0.23 \pm 0.17$ |

Table 10. Performance on simulated data, with $k = 2$ and non-normalized data.

| $k = 2$, Non-normalized | $\mu = 2$ $\sigma = 0$ | $\mu = 2$ $\sigma = 0.05$ | $\mu = 2$ $\sigma = 0.2$ | $\mu = 2$ $\sigma = 0.5$ | $\mu = 4$ $\sigma = 0$ | $\mu = 4$ $\sigma = 0.05$ | $\mu = 4$ $\sigma = 0.2$ | $\mu = 4$ $\sigma = 0.5$ |
|---|---|---|---|---|---|---|---|---|
| **FRIGATE** | $1 \pm 0$ | $1 \pm 0$ | $1 \pm 0$ | $1 \pm 0$ | $1 \pm 0$ | $1 \pm 0$ | $1 \pm 0$ | $1 \pm 0$ |
| **FRIGATE-MW** | $1 \pm 0$ | $1 \pm 0$ | $1 \pm 0$ | $1 \pm 0$ | $1 \pm 0$ | $1 \pm 0$ | $1 \pm 0$ | $1 \pm 0$ |
| **FRCM** | $1 \pm 0$ | $1 \pm 0$ | $1 \pm 0$ | $1 \pm 0$ | $1 \pm 0$ | $1 \pm 0$ | $1 \pm 0$ | $1 \pm 0$ |
| **FRSD** | $0.98 \pm 0.02$ | $0.99 \pm 0.02$ | $0.99 \pm 0.02$ | $1 \pm 0.02$ | $1 \pm 0$ | $1 \pm 0$ | $1 \pm 0$ | $1 \pm 0$ |
| **FRMV** | $0.12 \pm 0.21$ | $0.33 \pm 0.32$ | $0.05 \pm 0.1$ | $0.19 \pm 0.27$ | $0 \pm 0$ | $0 \pm 0$ | $0 \pm 0$ | $0 \pm 0$ |

Table 11. Performance on simulated data, with $k = 2$ and non-normalized data.

### 5. Supplementary 5 – Significance levels

P-values for the differences in performances of FRIGATE, FRIGATE-MW, FRMV, FRCM and FRSD of Datasets 5-7 in Table 2. The # column shows the ranking of ARI scores where 1 is best. In case of ties in the ARI we used the STD as a tiebreaker, where lower STD accounts for a better rank.

| | # | FRIGATE-MW | FRMV | FRSD | FRCM |
|---|---|---|---|---|---|
| FRIGATE | 2 | 0.256 | 4.66e-06 | 2.164e-07 | 3.764e-08 |
| FRIGATE-MW | 1 | - | 3.763e-09 | 1.308e-14 | 1.066e-15 |
| FRMV | 4 | - | - | 0.856 | 0.328 |
| FRSD | 3 | - | - | - | 3.611e-07 |
| FRCM | 5 | - | - | - | - |

Table S12. T-test p-values of ARI scores for ⌈25%⌉ features of Dataset 5.

| | # | FRIGATE-MW | FRMV | FRSD | FRCM |
|---|---|---|---|---|---|
| FRIGATE | 2 | 0.66 | 2.117e-12 | 1.771e-11 | 0.0961 |
| FRIGATE-MW | 1 | - | 2.031e-12 | 1.65e-11 | 0.0451 |
| FRMV | 5 | - | - | 0.029 | 5.446e-12 |
| FRSD | 4 | - | - | - | 5.575e-11 |
| FRCM | 3 | - | - | - | - |

Table S13. T-test p-values of ARI scores for ⌈50%⌉ features of Dataset 5.

| | # | FRIGATE-MW | FRMV | FRSD | FRCM |
|---|---|---|---|---|---|
| FRIGATE | 3 | 0.648 | 0.987 | 0.002 | 0.595 |
| FRIGATE-MW | 2 | - | 0.588 | 4.564e-05 | 1 |
| FRMV | 4 | - | - | 1.836e-04 | 0.504 |
| FRSD | 5 | - | - | - | 4.218e-08 |
| FRCM | 1 | - | - | - | - |

Table S14. T-test p-values of ARI scores for ⌈25%⌉ features of Dataset 6.

| | # | FRIGATE-MW | FRMV | FRSD | FRCM |
|---|---|---|---|---|---|
| FRIGATE | 2 | 0.231 | 0.607 | 0.021 | 0.127 |
| FRIGATE-MW | 4 | - | 0.653 | 0.052 | 3.07e-04 |
| FRMV | 3 | - | - | 0.057 | 0.052 |
| FRSD | 5 | - | - | - | 0.001 |
| FRCM | 1 | - | - | - | - |

Table S15. T-test p-values of ARI scores for ⌈50%⌉ features of Dataset 6.

| | # | FRIGATE-MW | FRMV | FRSD | FRCM |
|---|---|---|---|---|---|
| FRIGATE | 3 | 0.712 | 0.871 | 2.09e-05 | 0.241 |
| FRIGATE-MW | 2 | - | 0.209 | 4.39e-11 | 0.011 |
| FRMV | 4 | - | - | 4.173e-10 | 4.72e-04 |
| FRSD | 5 | - | - | - | 1.073e-13 |
| FRCM | 1 | - | - | - | - |

Table S16. T-test p-values of ARI scores for ⌈25%⌉ features of Dataset 7.

| | # | FRIGATE-MW | FRMV | FRSD | FRCM |
|---|---|---|---|---|---|
| FRIGATE | 1 | 0.177 | 0.018 | 1.232e-23 | 0.278 |
| FRIGATE-MW | 3 | - | 0.641 | 1.809e-15 | 0.76 |
| FRMV | 4 | - | - | 6.575e-18 | 0.394 |
| FRSD | 5 | - | - | - | 1.918e-16 |
| FRCM | 2 | - | - | - | - |

Table S17. T-test p-values of ARI scores for ⌈50%⌉ features of Dataset 7.

| | # | FRIGATE-MW | FRMV | FRSD | FRCM |
|---|---|---|---|---|---|
| FRIGATE | 2 | 0.526 | 1.031e-07 | 0.142 | 1.572e-18 |
| FRIGATE-MW | 3 | - | 1.66e-06 | 0.165 | 5.947e-12 |
| FRMV | 4 | - | - | 3.059e-08 | 0.468 |
| FRSD | 1 | - | - | - | 2.904e-22 |
| FRCM | 5 | - | - | - | - |

Table S18. T-test p-values of ARI scores for ⌈25%⌉ features of Dataset 8.

| | # | FRIGATE-MW | FRMV | FRSD | FRCM |
|---|---|---|---|---|---|
| FRIGATE | 1 | 0.15 | 3.753e-05 | 0.898 | 3.125e-04 |
| FRIGATE-MW | 3 | - | 7.479e-05 | 0.095 | 0.006 |
| FRMV | 5 | - | - | 3.53e-05 | 2.41e-04 |
| FRSD | 2 | - | - | - | 1.076e-05 |
| FRCM | 4 | - | - | - | - |

Table S19. T-test p-values of ARI scores for ⌈50%⌉ features of Dataset 8.

### 6. Supplementary 6 - Clinical significance

Full results of the ranked features by FRIGATE and FRIGATE-MW that were also top features found in [11]. See chapter 4 for details.

| Rank | Feature | Mendelson Cohen et al. |
|---|---|---|
| 1 | days in hospital | ☒ |
| 2 | Calculated Bicarbonate, Whole Blood | ☒ |
| 3 | Hemoglobin | ☑ |
| 4 | Hematocrit | ☑ |
| 5 | Platelet Count | ☑ |
| 6 | MCH | ☒ |
| 7 | Alanine Aminotransferase (ALT) | ☑ |
| 8 | Asparate Aminotransferase (AST) | ☒ |
| 9 | Bilirubin, Total | ☑ |
| 10 | Red Blood Cells | ☑ |
| 11 | Fibrinogen, Functional | ☒ |
| 12 | MCHC | ☒ |
| 13 | Calculated Total CO2 | ☒ |
| 14 | Anion Gap | ☒ |
| 15 | MCV | ☒ |
| 16 | Bicarbonate | ☒ |
| 17 | Alkaline Phosphatase | ☑ |
| 18 | Age | ☒ |
| 19 | pCO2 | ☒ |
| 20 | Neutrophils | ☒ |
| 21 | Lymphocytes | ☒ |
| 22 | pH | ☒ |
| 23 | Base Excess | ☒ |
| 24 | PT | ☒ |
| 25 | Glucose | ☒ |
| 26 | INR(PT) | ☒ |
| 27 | Chloride | ☒ |
| 28 | pO2 | ☒ |
| 29 | RDW | ☒ |
| 30 | Sodium, Whole Blood | ☒ |
| 31 | pH | ☒ |
| 32 | Ethanol | ☒ |
| 33 | Specific Gravity | ☒ |
| 34 | Creatinine | ☑ |
| 35 | Phosphate | ☒ |
| 36 | White Blood Cells | ☒ |
| 37 | Basophils | ☑ |
| 38 | Potassium, Whole Blood | ☒ |
| 39 | Urea Nitrogen | ☑ |
| 40 | Monocytes | ☑ |
| 41 | Calcium, Total | ☒ |
| 42 | Lactate | ☒ |
| 43 | Eosinophils | ☑ |
| 44 | PTT | ☒ |
| 45 | Magnesium | ☒ |
| 46 | Amylase | ☒ |
| 47 | Lipase | ☒ |

Table S20. Average ranks of FRIGATE-MW for Dataset 6. The bold line represents ⌈25%⌉ of features cutoff. ☑: Top features in [11], ☒: The rest.

| Rank | Feature | Mendelson Cohen et al. |
|---|---|---|
| 1 | days in hospital | ☒ |
| 2 | Platelet Count | ☑ |
| 3 | Calculated Bicarbonate, Whole Blood | ☒ |
| 4 | Hematocrit | ☑ |
| 5 | pH | ☒ |
| 6 | MCH | ☒ |
| 7 | Age | ☒ |
| 8 | Hemoglobin | ☑ |
| 9 | Neutrophils | ☒ |
| 10 | Red Blood Cells | ☑ |
| 11 | INR(PT) | ☒ |
| 12 | Bilirubin, Total | ☑ |
| 13 | Alanine Aminotransferase (ALT) | ☑ |
| 14 | Asparate Aminotransferase (AST) | ☒ |
| 15 | pH | ☒ |
| 16 | Lymphocytes | ☒ |
| 17 | Fibrinogen, Functional | ☒ |
| 18 | MCHC | ☒ |
| 19 | MCV | ☒ |
| 20 | Anion Gap | ☒ |
| 21 | Calculated Total CO2 | ☒ |
| 22 | Alkaline Phosphatase | ☑ |
| 23 | RDW | ☒ |
| 24 | PT | ☒ |
| 25 | Glucose | ☒ |
| 26 | Specific Gravity | ☒ |
| 27 | Urea Nitrogen | ☑ |
| 28 | Bicarbonate | ☒ |
| 29 | pCO2 | ☒ |
| 30 | Basophils | ☑ |
| 31 | Base Excess | ☒ |
| 32 | Ethanol | ☒ |
| 33 | White Blood Cells | ☒ |
| 34 | Chloride | ☒ |
| 35 | Creatinine | ☑ |
| 36 | pO2 | ☒ |
| 37 | Sodium, Whole Blood | ☒ |
| 38 | Potassium, Whole Blood | ☒ |
| 39 | Monocytes | ☑ |
| 40 | Eosinophils | ☑ |
| 41 | Phosphate | ☒ |
| 42 | PTT | ☒ |
| 43 | Calcium, Total | ☒ |
| 44 | Magnesium | ☒ |
| 45 | Lactate | ☒ |
| 46 | Amylase | ☒ |
| 47 | Lipase | ☒ |

Table S21. Average ranks of FRIGATE for Dataset 6. The bold line represents ⌈25%⌉ of features cutoff. ☑: Top features in [11], ☒: The rest.

### 7. Supplementary 7 – The effect of $k$ in each algorithm

FRIGATE requires determining the number of clusters $k$ in advance, unlike FRCM and FRSD, which test a prescribed range of $k$. Generally, it is preferred not to determine the number of clusters in advance. However, all the algorithms mentioned above are based on k-means, which requires $k$ as an input. As is commonly done, we suggest trying different $k$ values and choosing the value using some criterion, e.g., the "elbow" method [8]. This is the approach we use for FRIGATE.

To test the effect of $k$ in prior algorithms, we simulated data with four clusters ($k = 4$), fifty samples in each cluster ($c = 50$), and 100 features, of which 20 are informative ($\alpha = 20$, $\beta = 80$), when the data is z-score normalized, and measures the accurate recognition rate. We repeated the run 10 times for each case and present in Figure S11 the mean results. We ran FRCM and FRSD with a range $[min\_k, max\_k]$ where $min\_k$ of {2,3,4}, and $max\_k = \{4,8,12,15\}$, (recall that for FRCM and FRSD the default range is [2:15] as $k\_max = \min(\lceil\sqrt{200}\rceil, 20) = 15$). We can see that generally an increase of $max\_k$ results in a sharp decrease in the results of FRSD. Interestingly, using $min\_k < k$ had a positive effect: using a range of [2:4], when $k = 4$, produced better results than using [3,4] or [4,4]. Similar results were obtained for simulation with $\mu = 4$ (Figure S13).

It is worth mentioning that in FRSD the larger the range - the more iterations the algorithm performs in total ($T = 200$ for each tested value of $k$). To see if the total number of iterations has a major effect on the results, we also tested the performance of FRSD when the range $[k, k]$ was given as input for different $k$ values, and the total number of iterations per $k$ was set to 2800 (the number of iterations for the full default range of $k$), and the results are shown in Figure S12 for $\mu = 2$ (see Figure S14 for $\mu = 4$). We can see that FRSD performs best for k=3 and k=2, although the real number of clusters is four. Higher values of $k$ harm the ability of the algorithm to find any relevant features. This helps explaining the results in Figure S11, where we see a drop in performance when large values of $k$ are included and affecting the results.

FRCM, on the other hand, has a fixed number of iterations and samples a value of $k$ for each iteration from the same range as FRSD. Interestingly, when performing the same analyses for FRCM, the algorithm produced perfect results for all cases (results not shown).

Unlike FRSD and FRCM, FRIGATE and FRMV require $k$ as an input. Hence, we tested how different values of input $k$ affect the results of FRIGATE and FRMV on simulated data with the same parameters used for FRSD and FRCM. We tested the algorithms in the same simulation, using as input values of $k$ {2, 3, 4, 8,

12, 15}. The results of FRIAGTE were perfect in all scenarios, which means it was unaffected by the input $k$. FRMV had poor results in all scenarios (results not shown).

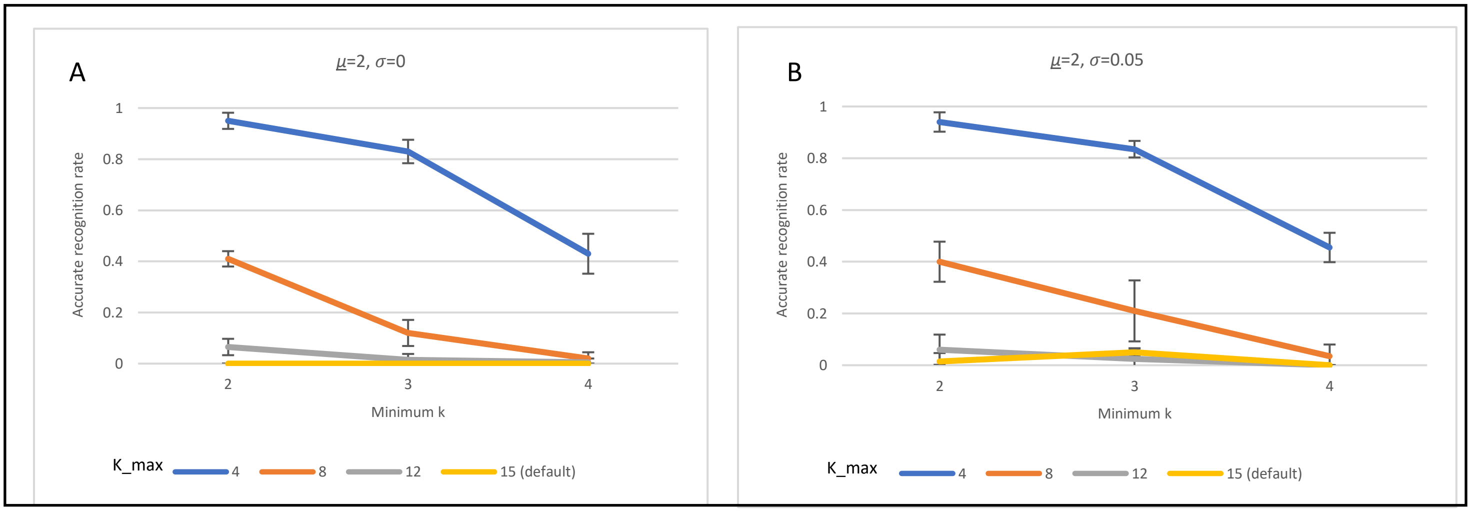

Figure S11. Performance of FRSD with different ranges for $[k\_min, k\_max]$. The simulation parameters are 4 clusters, 50 samples in each cluster, 100 features - of which 20 are informative, $\mu = 2$. The results show accurate recognition rate of FRSD over 10 runs. A: $\sigma = 0$, B: $\sigma = 0.05$.

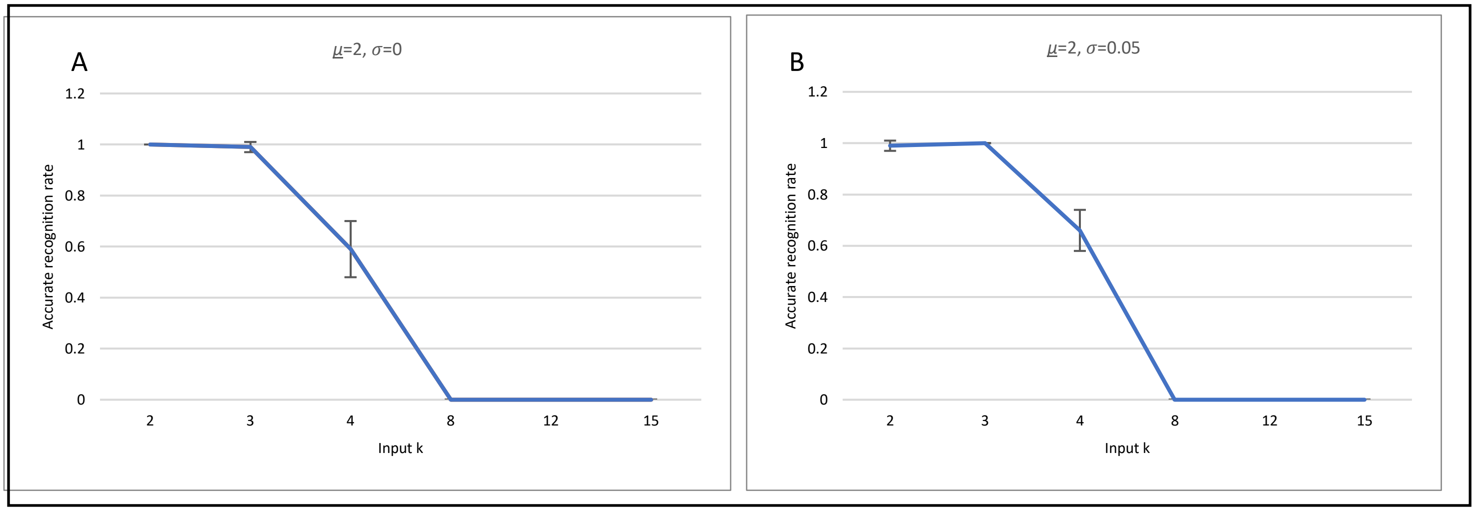

Figure S12. Performance of FRSD when a single number of clusters is tested, the true number is 4, and 2800 iterations are performed per each number of clusters tested. The rest of the simulation parameters are as in Figure 7. A: $\sigma = 0$, B: $\sigma = 0.05$.

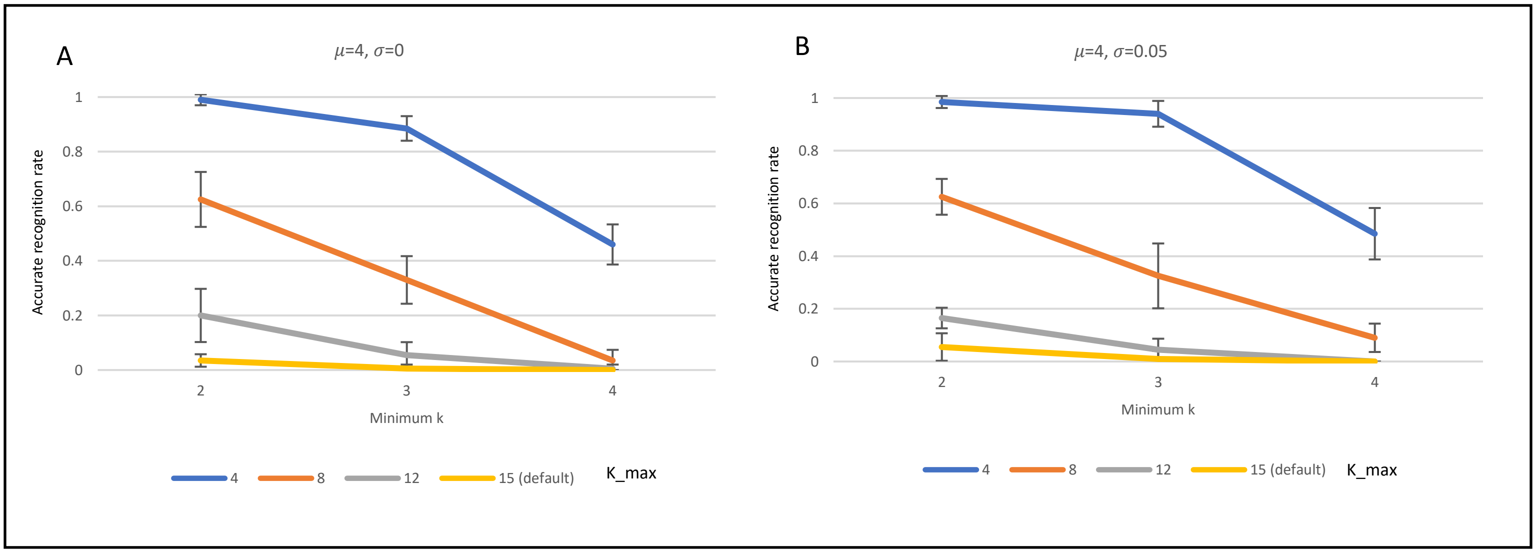

Figure S13. Performance of FRSD with different values used as $max\_x$. The simulation has 4 clusters, 50 samples in each cluster, 100 features - 20 of which are informative for clustering, $\mu = 4$ and $\sigma = 0$ or $\sigma = 0.05$ for A and B respectively. We measured the accurate recognition rate and we present here the mean of 10 runs of FRSD on the range $[min\_k, max\_k]$ for different values of min_x and max_x.

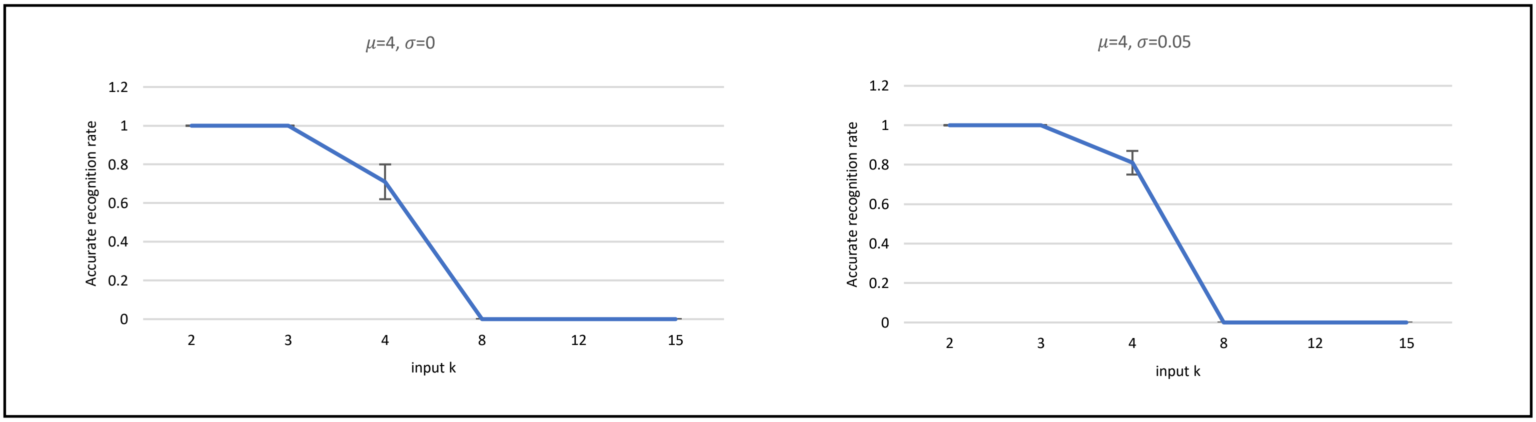

Figure S14. Performance of FRSD with a single value of $k$ and 2800 iterations. The simulation parameters are as in Figure S6. A: $\sigma = 0$, B: $\sigma = 0.05$.
